# Contemporary syphilis is characterised by rapid global spread of pandemic *Treponema pallidum* lineages

**DOI:** 10.1101/2021.03.25.21250180

**Authors:** Mathew A. Beale, Michael Marks, Michelle J. Cole, Min-Kuang Lee, Rachel Pitt, Christopher Ruis, Eszter Balla, Tania Crucitti, Michael Ewens, Candela Fernández-Naval, Anna Grankvist, Malcolm Guiver, Chris R. Kenyon, Rafil Khairulin, Ranmini Kularatne, Maider Arando, Barbara J. Molini, Andrey Obukhov, Emma E. Page, Fruzsina Petrovay, Cornelis Rietmeijer, Dominic Rowley, Sandy Shokoples, Erasmus Smit, Emma L. Sweeney, George Taiaroa, Jaime H. Vera, Christine Wennerås, David M. Whiley, Deborah A. Williamson, Gwenda Hughes, Prenilla Naidu, Magnus Unemo, Mel Krajden, Sheila A. Lukehart, Muhammad G. Morshed, Helen Fifer, Nicholas R. Thomson

## Abstract

Syphilis is an important sexually transmitted infection caused by the bacterium *Treponema pallidum* subspecies *pallidum*. The last two decades have seen syphilis incidence rise in many high-income countries, yet the evolutionary and epidemiological relationships that underpin this are poorly understood, as is the global *T. pallidum* population structure. We assembled a geographically and temporally diverse collection of clinical and laboratory samples comprising 726 *T. pallidum* genomes. We used detailed phylogenetic analysis and clustering to show that syphilis globally can be described by only two deeply branching lineages, Nichols and SS14. We show that both of these lineages can be found circulating concurrently in 12 of the 23 countries sampled. To provide further phylodynamic resolution we subdivided *Treponema pallidum* subspecies *pallidum* into 17 distinct sublineages. Importantly, like SS14, we provide evidence that two Nichols sublineages have expanded clonally across 9 countries contemporaneously with SS14. Moreover, pairwise genome analysis showed that recent isolates circulating in 14 different countries were genetically identical in their core genome to those from other countries, suggesting frequent exchange through international transmission pathways. This contrasts with the majority of samples collected prior to 1983, which are phylogenetically distinct from these more recently isolated sublineages. Bayesian temporal analysis provided evidence of a population bottleneck and decline occurring during the late 1990s, followed by a rapid population expansion a decade later. This was driven by the dominant *T. pallidum* sublineages circulating today, many of which are resistant to macrolides. Combined we show that the population of contemporary syphilis in high-income countries has undergone a recent and rapid global expansion.

## Introduction

Syphilis, caused by the bacterium *Treponema pallidum* subspecies *pallidum* (TPA), is a prevalent sexually transmitted infection which can cause severe long-term sequelae when left untreated. Historically, syphilis is commonly believed to have caused a large epidemic across Renaissance Europe, having previously been absent or unrecognised^1,2^. Although accurate dating of the most recent common ancestor of TPA is still the subject of debate^3–6^, it is suggested that the strains of TPA that persist in the human population to this day can be traced back to that introduction into Western Europe approximately 500 years ago, and subsequently disseminated globally^3,4,6^.

Following the introduction of effective antibiotics after World War II, syphilis incidence fluctuated^7^ without disappearing, until the 1980s and 1990s during the HIV/AIDS crisis when disease incidence declined markedly^8^, linked to community wide changes in sexual behaviour, shifting of affected populations, AIDS-related mortality and widespread antimicrobial prophylaxis of HIV infected populations. However, since the beginning of the 21^st^ century, there has been a substantial resurgence in syphilis incidence^9–13^. In many countries, this has been associated with populations of men who have sex with men (MSM) engaging in high risk sexual activity^11,14^. Transmission between MSM and heterosexuals is a particular concern due to the risk of *in utero* transmission to the foetus, leading to congenital syphilis^15^.

Previous genomic analyses of TPA genomes have described two deep branching phylogenetic lineages ‘SS14’ and ‘Nichols’^3^. SS14-lineage strains represent the vast majority of published genomes^4^, and phylogenetic analysis showed that the origins of the SS14-lineage can be traced back to the 1950s^3^, followed by subsequent expansion of sublineages occurring during the 1990s^4^. Our understanding of Nichols-lineage is predominantly limited to laboratory strains from the USA, with relatively few clinical strains sequenced^4,16^. However, most TPA genomes published to date originate from the USA^4^, Western Europe^3,4,16,17^ and China^18,19^, and our understanding of the true breadth of diversity of TPA is incomplete^20^. This is partly explained, by the fact it has not been possible to culture TPA outside of a rabbit until recently^21^, and also explains why our view of the diversity of syphilis samples predating the 21^st^ century is even more limited.

In this multicentre collaborative study, we used direct whole genome sequencing to generate a global view of contemporary syphilis from patients in Africa, Asia, the Caribbean, South America and Australia sampled between 1951 and 2019. Our dataset also includes a detailed analysis of the ‘within-country’ variation seen in TPA genomes in North America and Europe. We present evidence of globally spanning transmission networks with identical strains found in dispersed countries, indicating that, based on our data, TPA is essentially panmictic. Furthermore, we show that this genetic homogeneity is the result of a rapid and global expansion of TPA sublineages occurring within the last 30 years following a population bottleneck. This means the TPA population infecting patients in the 21^st^ century is not the same as that infecting patients in the 20^th^.

## Results

### Describing the global population structure of *Treponema pallidum* subspecies *pallidum*

We performed targeted sequence-capture whole genome sequencing on residual genomic DNA extracted from diagnostic swabs taken from TPA PCR positive syphilis patients, and on TPA strains previously isolated in rabbits. We combined these data with 133 previously published genomes^3,4,17–19,22–25^. After assessment for genome coverage and quality, we had a total of 726 genomes with >25% of genome positions at >5X coverage (mean 82%, range 25-97%), sufficient for primary lineage classification. This dataset included 577 new genomes sequenced directly from clinical samples, and 16 new genomes sequenced from samples passaged in rabbits.

Our dataset includes 23 countries (range 1-355 genomes per country; Figure 1A, 1C), including previously poorly or unsampled regions including Africa (South Africa (n=1) and Zimbabwe (n=18)), Scandinavia (Sweden, n=7), Central Europe (Hungary, n=20), Central Asia (Tuva Republic, Russia, n=10) and Australia (n=5), as well as substantially increasing the sampling from North America (Canada, n=157; United States, n=86) and Western Europe (Spain, n=5; Belgium, n=1; Ireland, n=4; the United Kingdom, n=355). Due to a lack of long term archived samples, 96.0% (n=697) of samples were collected from 2000 onwards (Figure 1B, 1E). Samples collected prior to 2000 (n=29) were passaged in the in vivo rabbit model (Supplementary Data 1), whereas most samples collected from 2000 onwards (89.8%, 626/697) were sequenced directly from clinical samples.

**Figure 1.**
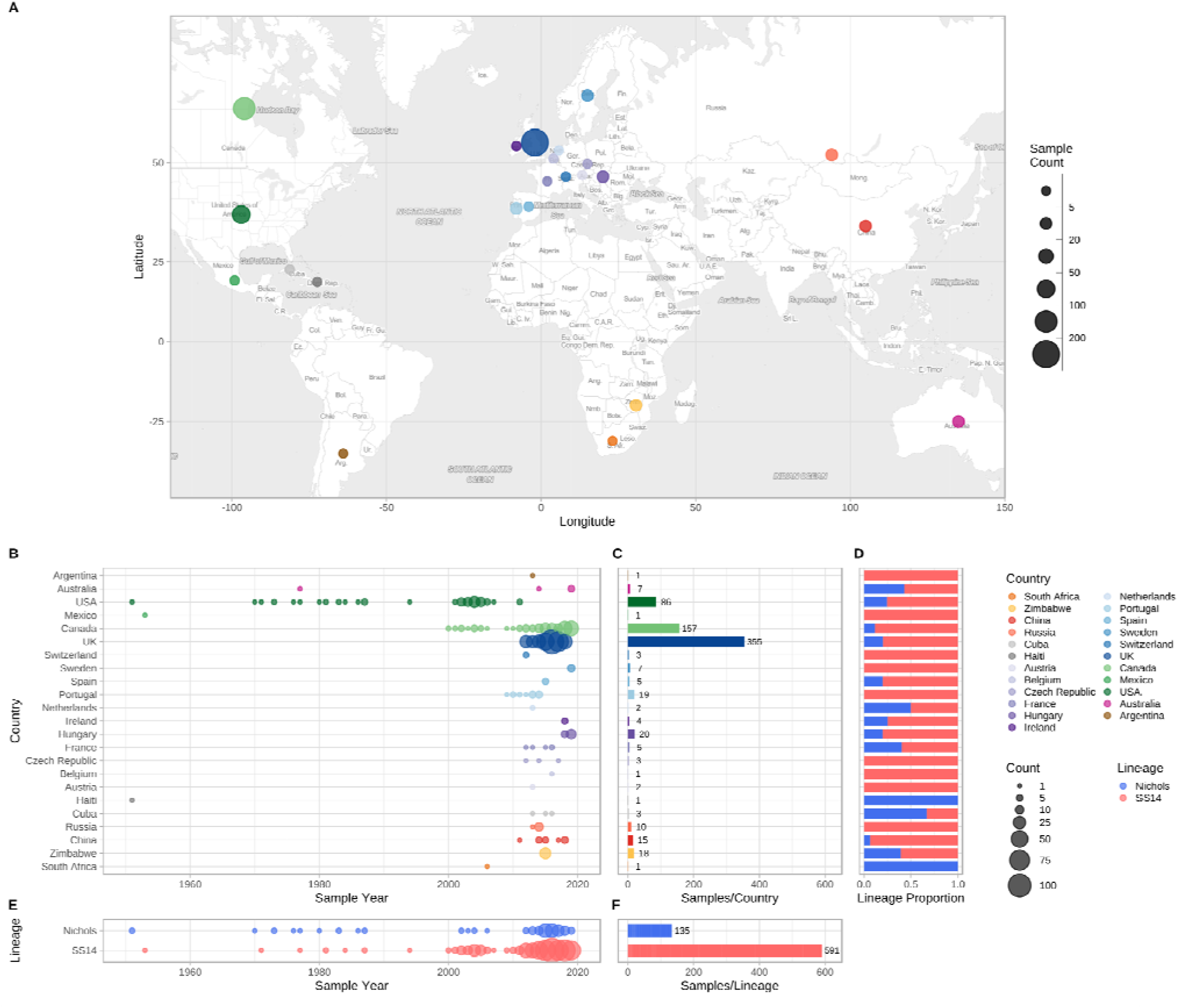
Global distribution of 726 *Treponema pallidum* subspecies *pallidum* partial genomes. A-Map of sampled countries for 726 partial (>25% of genome positions) genomes. Circle size corresponds to total number of genomes (binned into categories), and colour corresponds to country. Country coordinates used are the country centroid position, apart from Russia (where the centroid for the Tuva Republic is used) and Mexico (where the location of Mexico City is used). Map tiles by Stamen Design (CC-BY 3.0), map data by OpenStreetMap (ODbl). B-Temporal distribution of samples by country. Size of circle indicates number of samples for that year. Three samples from Baltimore (USA) had uncertain sampling dates (1960-1980) and were set to 1970 for plotting dates (1B, 1E). Genomes derived from passaged variants of the Nichols-1912 isolate or with uncertain collection dates are not shown in plotted timeline (1B, 1E). C-Total count of samples by country. D-Relative proportion of country samples corresponding to each TPA lineage (where only one sample was present per country, shows the lineage this corresponds to). E-Temporal distribution of the samples by TPA lineage. F-Total count of samples by TPA Lineage.

Phylogenetic analysis assigned all genomes into one of two deeply branching lineages (“Nichols” or “SS14”) (Supplementary Figure 1). Looking across all well sampled countries (Figure 1C, 1D), from the first detection of the modern SS14-lineage (excluding the outlying 1953 Mexico A strain) in the 1970s, we consistently see both lineages circulating broadly through until 2019 (Figure 1B, 1E). More specifically 81.3% (n=590) of genomes belonged to the SS14-lineage and 18.7% (n=136) to the Nichols-lineage, and in the 12/23 countries where both lineages were present, 80.3% (544/677, median per country 75.3%, range 33.3-93.3%) were SS14-lineage (Figure 1D, 1F). In the 11 countries showing only a single lineage, most had three or fewer samples, the notable exceptions being Portugal (n=19), Sweden (n=7) and Russia (n=10) (Figure 1C).

### Fine scaled analysis of SS14- and Nichols-lineage phylogenies

To answer finer scaled evolutionary questions, we filtered our dataset to focus on the 528 genomes with >75% genome sites at >5X (genome length 1,139,569 bp, mean % sites 92.9%, range 75.1–96.9%) and a mean coverage of 111X (range 11X–727X). This filtered dataset comprised 401 new and 127 published genomes (Supplementary Data 1), but excluded the only sample from Belgium, leaving 22 countries in the analysis. After excluding 19 regions of recombination and genomic uncertainty due to gene orthology or repetitive regions^3,4,6^, we used Gubbins^26^ to infer a further 19 regions of putative recombination (see Methods and Supplementary Data 2 for details). We refer to the resulting masked sequence alignment as the core genome and used it to infer a whole genome maximum likelihood phylogeny using IQ-Tree^27^ (Supplementary Figure 2). To define sublineage clusters we used 100 bootstrapped trees as independent inputs to rPinecone^28^ with a 10 SNP threshold, and evaluated their consistency using hierarchical clustering (Supplementary Figure 3). We found broad support for the Nichols sublineages across all conditions evaluated, but some parts of the SS14-lineage phylogeny were less well supported. To focus on the more stable sublineages, we required that at least 5% of the bootstrap replicates supported a cluster. Using this approach, we defined 17 sublineages and 8 singleton strains across both SS14-and Nichols-lineages (SS14: 426 genomes divided into 5 sublineages and 4 singletons; Nichols: 102 genomes divided into 12 sublineages and 4 singletons; Figure 2; Supplementary Figures 2, 4, 5).

**Figure 2.**
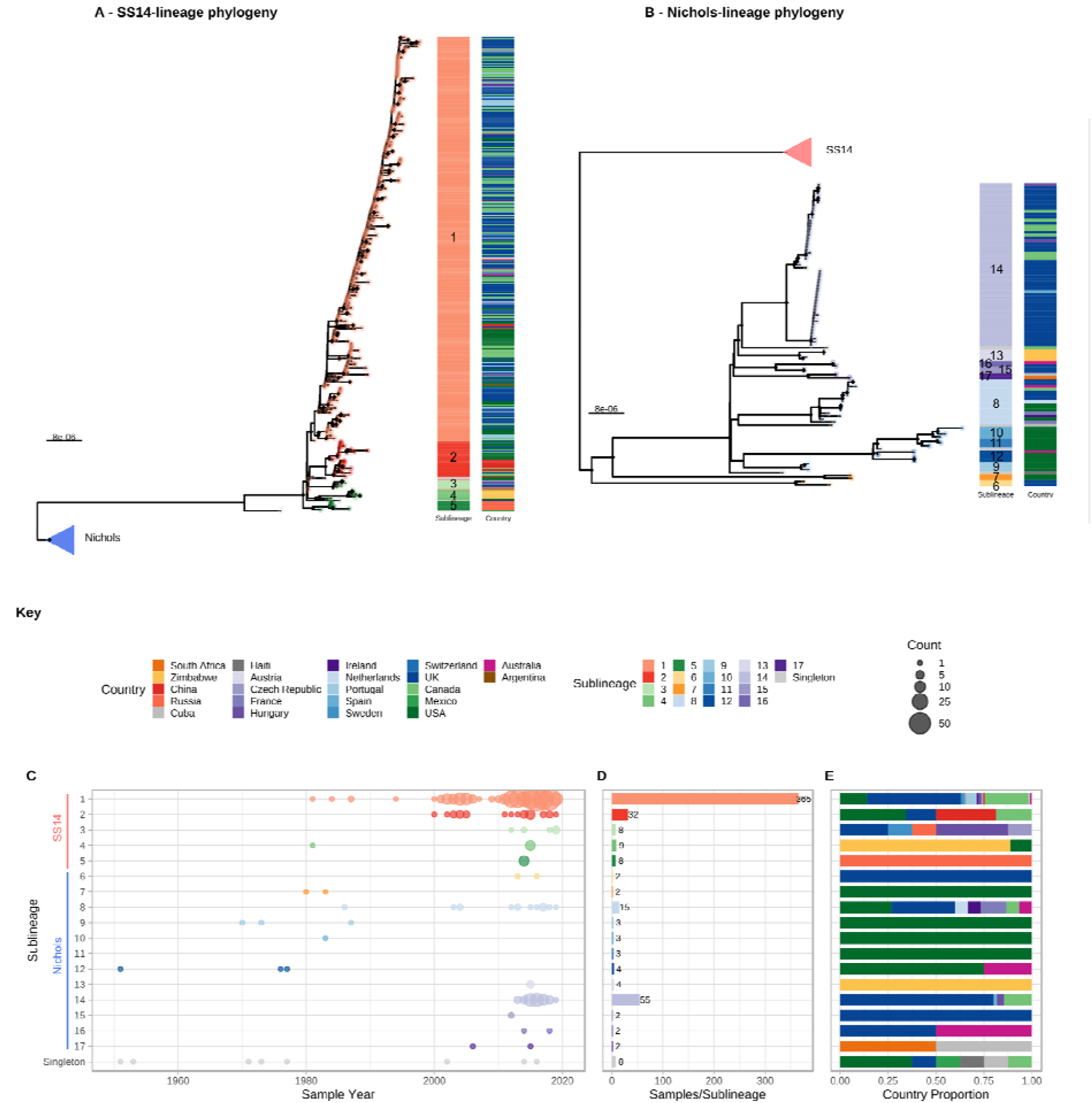
Fine scaled analysis of 528 high quality (>75% reference sites) TPA genomes and sublineages. A-Recombination masked WGS phylogeny, highlighting the SS14-lineage (n=426). B-Recombination masked WGS phylogeny, highlighting the Nichols-lineage (n=102), including four outlying genomes (sublineages 6 & 7). For A and B, coloured strips show sublineage and country; Tree tips show sublineage. Coloured triangle indicates node position of collapsed sister lineage. UF Bootstraps >=95% are marked with black node marks. C-Temporal distribution of samples by sublineage (unrelated singleton genomes are grouped together). D-Total count of samples by sublineage. E-Relative proportion of each sublineage samples corresponding to country.

From Figure 2 it is apparent that the phylogeny of SS14-lineage is dominated by SS14 sublineage 1 (n=365), comprised of closely related genomes present in 18 countries and six continental groupings (Asia, Caribbean, Europe, North America, Oceania, and South America). The oldest example of sublineage 1 was collected in 1981 (TPA_USL-SEA-81-3, Seattle USA), and the most recent samples were from 2019 (Figure 2C). Sublineage 2 (n=32; Figure 2E, Supplementary Figure 4) contained samples from Canada, China, the UK and the USA. In a previous analysis^4^, we manually divided this sublineage into two groups (one from China, one from the USA), based on temporal and geospatial divergence, and the independent evolution of different macrolide resistance alleles. However, by adding new genomes (Supplementary Figure 4), we now see that even the original cluster of samples from China is interspersed with genomes from the UK (n=1) and Canada (n=4), indicating that this is not a geographically restricted group.

The 12 2015 Zimbabwean genomes in our study formed two distinct clades, one nested within SS14-lineage (sublineage 4, n=8, also containing a single distantly related sample from the USA 1981, TPA_USL-SEA-81-8), the other within Nichols-lineage (sublineage 13, n=4) and exclusively found in Zimbabwe. We also examined 10 genomes collected in the Tuva Republic, central Russia in 2013/2014, and these were distributed between three different SS14 sublineages (1, 3, 5). Sublineage 5 was found only in Tuva, whilst sublineage 3 was found throughout Europe (Czech Republic, Hungary, Sweden and the UK; Figure 2E, Supplementary Figure 4), with the remaining sample from Russia belonging to the highly expanded global SS14 sublineage 1.

Consistent with previous studies^3,4,16^ Nichols-lineage strains were genetically more diverse, with longer branch lengths and higher nucleotide diversity than SS14-lineage (Nichols π=3.2×10^−5^, SS14 π=6×10^−6^), reflecting the predicted age of lineage diversification. However, our increased sampling also revealed two recent clonal expansions within the Nichols-lineage: sublineage 14 (n=55), comprising samples from Canada, Hungary, Spain and the UK (Figure 2E) and sublineage 8 (n=15) comprising samples from the Australia, Canada, France, Ireland, the Netherlands, the UK, and the USA (Supplementary Figure 5).

In addition to observing evidence of recent clonal expansions we also show greater resolution of Nichols-lineage diversity, identifying two new samples from UK patients (PHE130048A, PHE160283A, collected in 2013 and 2016 respectively) which occupy positions basal to all Nichols-lineage strains (Supplementary Figure 5). Indeed, this analysis suggested the most recent common ancestor of this sublineage was very close to the root of all TPA.

Multiple derivatives of the highly passaged prototype Nichols-lineage strain (denoted Nichols-1912) isolated in 1912 were included in our analysis (Nichols_v2, Seattle_Nichols, Nichols_Houston_E, Nichols_Houston_J, and Nichols_Houston_O). Figure 2B (Supplementary Figure 6) shows that derivatives of Nichols-1912 fall within a distinct clade which also includes independently collected contemporaneous clinical samples (some with minimal passage through the rabbit model), including a sample (TPA_AUSMELT-1) collected in Australia in 1977^29^. This clade could be subdivided into four sublineages (9, 10, 11, 12) and one singleton. Notably, the last sample belonging to this clade was collected in 1987 (TPA_USL-Phil-3). Hence, within our sampling framework this appears to be an example of a historic lineage becoming extinct. More broadly, we note that although this cluster of both clinical and laboratory strains were all passaged to varying degrees through the rabbit, other samples passaged in the rabbit were distributed throughout the phylogeny and were present in 9/17 sublineages (Supplementary Figure 7).

### Temporal analysis of population dispersal

To infer temporal patterns within the global phylogeny, we performed Bayesian phylogenetic reconstruction using BEAST^30^ under a Strict Clock model with a Bayesian Skyline population distribution. We excluded 8 samples from strains known to be heavily passaged or with uncertain collection dates from the previous dataset of 528, giving a dataset of 520 samples and 883 variable sites. We inferred a median molecular clock rate of 1.28 ×10^−7^ substitutions/site/year (95% Highest Posterior Density (HPD) 1.07 ×10^−7^ – 1.48 ×10^−7^), equivalent to one substitution/genome every 6.9 years, consistent with recent analyses^4,6^.

Within the global TPA phylogeny (Figure 3A), we observed several patterns of genomic dispersal. The first reflects the separation of the Nichols- and SS14-lineages (median date in our analysis 1534, 95% Highest Posterior Density 1430-1621), the subject of much previous analysis^3,4,6^. These data also showed that the common ancestor of these lineages is separated from recent samples by long phylogenetic branches and an absence of older ancestral nodes, suggesting unsampled historical diversity, and that most contemporary TPA descend from much more recent ancestral nodes. We previously dated the common ancestors for clonal expansions of 9 SS14 sublineages between the 1980s and early 2000s^4^. With this expanded dataset, we focussed on the ancestral nodes leading to the major clonally expanded sublineages 1, 2, 8 and 14, each having at least 15 samples.

**Figure 3.**
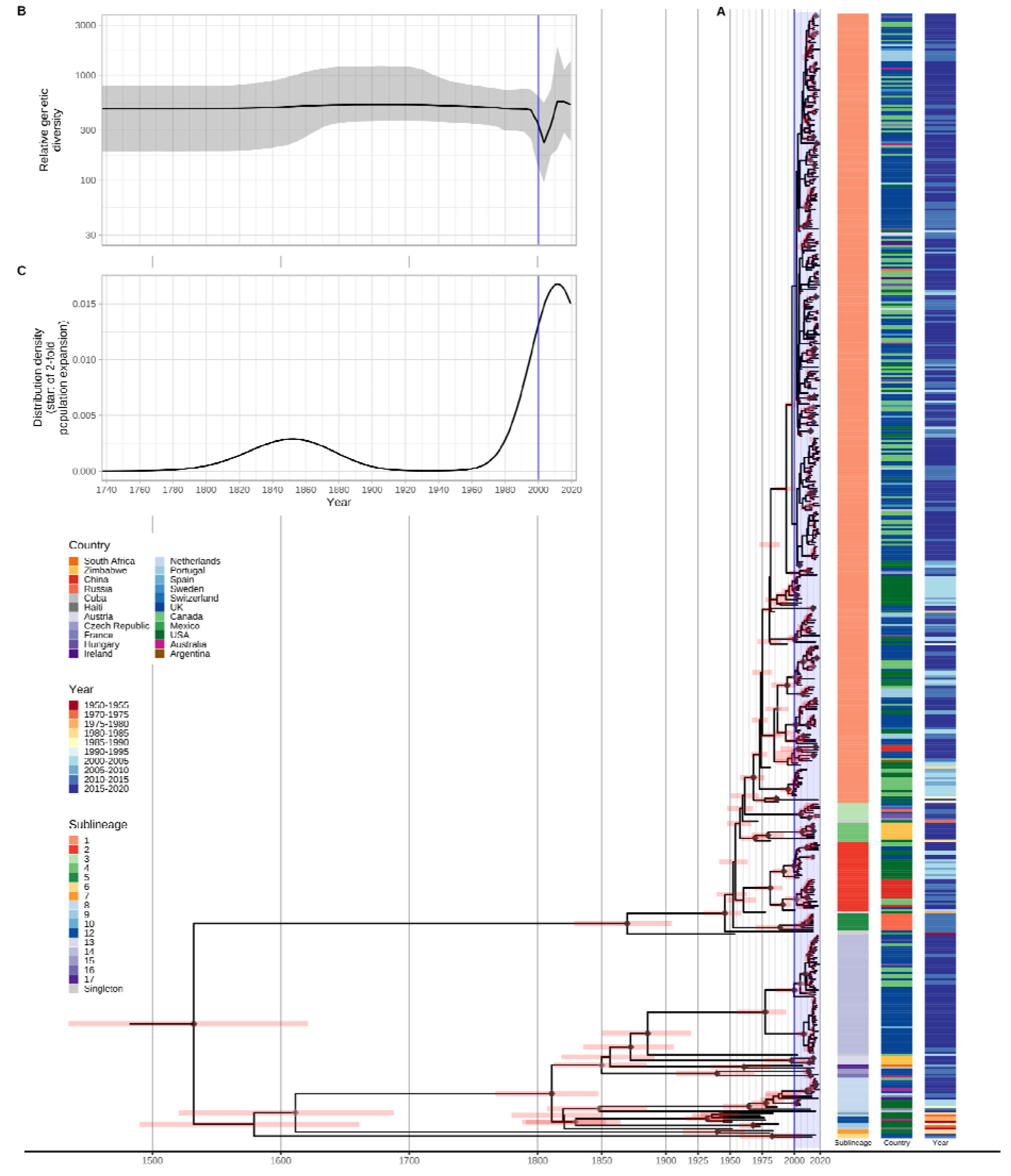
Bayesian maximum credibility phylogeny of 520 TPA genomes shows population contraction during the 1990s, followed by rapid expansion from the early 2000s onwards. A-Time-scaled phylogeny of 520 TPA genomes. Node points are shaded according to posterior support (black >96%, dark grey >91%, light grey >80%). Red bars on nodes indicate 95% Highest Posterior Density intervals. Blue line and shaded area highlight post-2000 expansion of lineages. B-Bayesian Skyline plot of genetic diversity shows small population expansion and contractions during the 19th and 20th Centuries, followed by a sharp decline and rapid re-emergence during the 1990s and 2000s. C-Posterior distribution of start dates for a 2-fold expansion above skyline mean shows strong support for expansion after 1990 in 68.6% (9263/13503) of trees.

Next, we used Bayesian Skyline analysis to determine the relative genetic diversity over different time periods in the phylogeny (Figure 3B), showing a very sharp decline during the 1990s and 2000s, followed by an equally sharp rise that continued until present day. To test the statistical support for this expansion, we extracted the proportion of trees in the posterior distribution supporting a >2-fold population expansion above the population mean (68.6%, 9263/13503 trees), and plotted the distribution of expansion start dates (Figure 3C) (median date 2011). We further tested the proportion of trees supporting a 2-fold population decline between 1990-2015 (90.7%, 12248/13503 trees, median date 2000) and a 2-fold population expansion between 1990-2015 (59.0%, 7966/13503 trees, median date 2012) (Supplementary Figure 8). These findings were also apparent in SS14 sublineages 1 and 2 (Supplementary 9) but not for Nichols sublineage 8. We had insufficient temporal signal to repeat this analysis on multi-country expanded Nichols sublineage 14 (Supplementary Figure 10).

### Global sharing of sublineages and identical strains

To further understand the patterns of recent population expansion we sought evidence of sharing of sublineages among countries, classifying sublineages as singletons (n=8), private to a country (n=8), or multi-country (n=9), and found that 20/22 countries contained at least one multi-country sublineage (Figure 4A, Supplementary Figure 11). We inferred pairwise SNP distances for genomes within and between each country (Figure 4C, 4D), and where there was more than one sample (n=18), we found fewer than 26 (SS14-lineage) and 80 (Nichols-lineage) pairwise SNPs separating genomes within any one country (Figure 4C), illustrating the close genetic relationships between samples (particularly SS14-lineage). We also found very low genetic distance between paired samples from different countries, with 27 country pairings (14 countries) showing zero core genome pairwise SNPs (Figure 4D). In particular Canada, UK and USA, with the highest sampling (Figure 4B), showed the most zero pairwise interactions with other countries (Figure 4D). Therefore, we cannot rule out similar transmission events occurring between other countries. We compared pairwise SNP distances with geographical distance between country centroids. Although we found a moderate correlation for Nichols-lineage (Pearson’s correlation 0.49, p<0.001), this was lower for SS14-lineage (0.31, p<0.001) and for the four largest multi-country sublineages (sublineage 1, 0.09, p<0.001; sublineage 2, 0.43, p<0.001; sublineage 8, 0.27, p<0.001; sublineage 14, 0.08, p<0.001) (Supplementary Figure 12). Hence, overall this indicates weak geographical structure for TPA.

**Figure 4.**
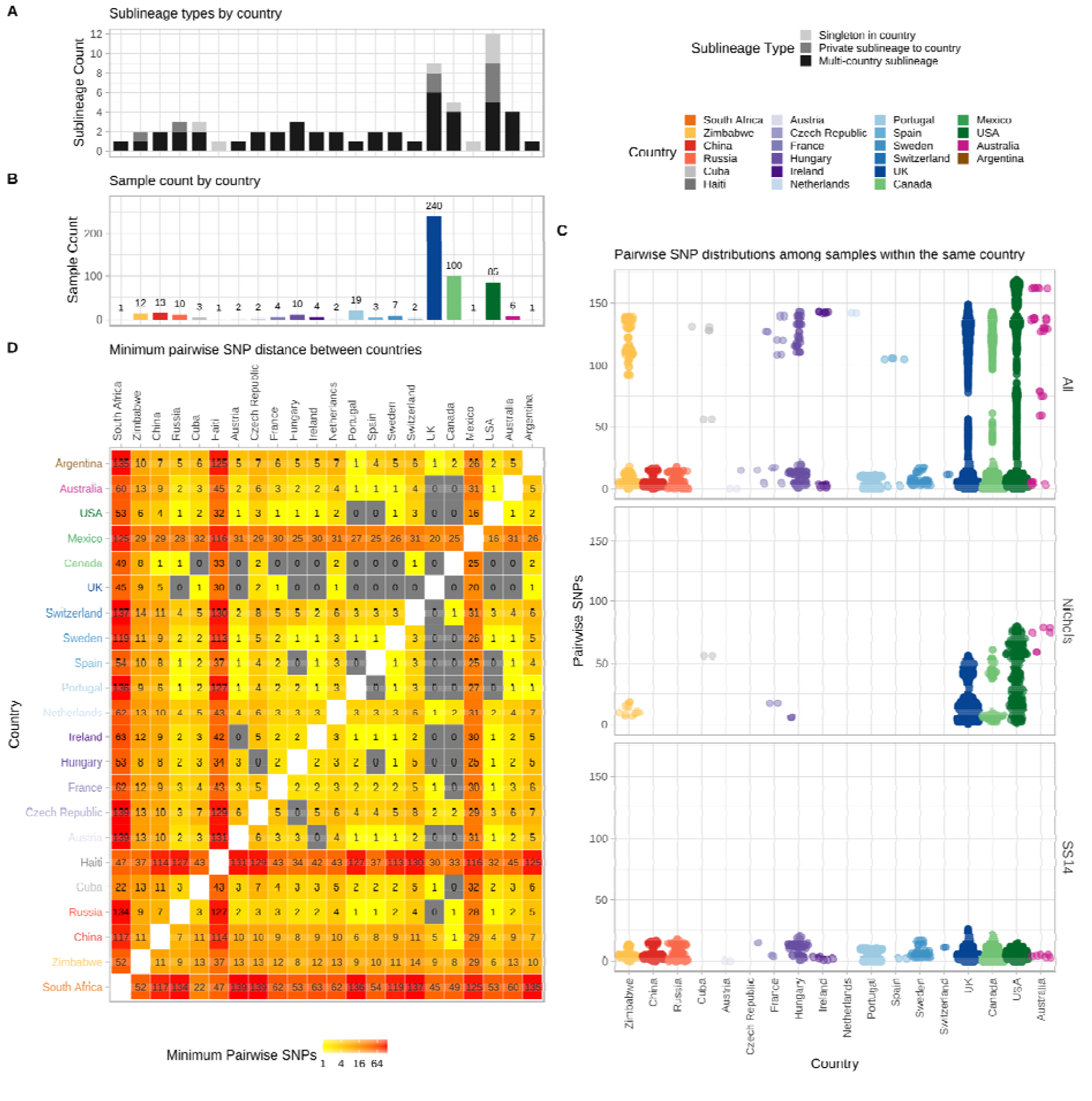
Substantial sharing of closely related strains within and between countries. A-Number of sublineages found per country, classified by sublineage distribution (multi-country=black, private to one country=medium grey, singleton=light grey). B-Total high-quality genomes per country. C-Pairwise comparison of SNP distance distributions from samples in each country (where >1 sample), across all samples and within lineages. D-Minimum pairwise SNPs between samples from different countries. All pairwise SNP comparisons exclude comparisons with same samples. Haiti, South Africa and Mexico appear striking outliers in terms of genetic relatedness (D), but this reflects that the Haiti and Mexico samples were collected in the 1950s, and we had only a single genome from these countries.

To understand these observations more fully, we focussed on British Columbia (Canada; BC) and England, both of which have experienced a recent rise in syphilis incidence (Supplementary Figure 13A), and for which we had a large number of samples. Included were 84 high quality BC genomes collected by the BC Centre for Disease Control between 2000 and 2018. From England, we had 240 high quality genomes from samples taken by the National Reference Laboratory at Public Health England (n=198) and non-referring laboratories in London (n=26), Brighton (n=9), Leeds (n=2) and Manchester (n=5) collected between 2012 and 2018. In BC, SS14 sublineage 1 dominated throughout the 18-year survey period, representing 82% of all BC genomes (Supplementary Figure 13B). In addition, isolated cases of SS14 sublineage 2 were seen in 2000 and 2012 as well as a single Nichols-lineage sample (singleton) in 2002 (Supplementary Figure 13B). Then from 2013 onwards, we detected two new Nichols sublineages: Nichols sublineage 8 and sublineage 14. The latter two lineages were also found across USA and Europe (Figure 2E).

Both Nichols- and SS14-lineages were consistently present in the English samples between 2012 and 2018. All of the common sublineages (4/4) found in BC were also present in England, as well as 4 additional sublineages (Nichols sublineages 6, 15, 16; SS14 sublineage 3) and one SS14 singleton strain not detected in BC (Supplementary Figure 13B). Sublineage 14, first detected in BC in 2013, was also the most numerous Nichols sublineage in England, but notably was not detected here until 2014.

Pairwise SNP distances between orthogonal genomes from the same sublineages showed 2622 pairwise combinations of BC (n=56) and English samples (n=78) sharing zero pairwise SNPs over the core genome alignment for isolates collected between 2004 and 2019. To understand the effect of temporal distance we compared both the pairwise SNP distance and the pairwise time distance (in years) between genomes from the same sublineage (Supplementary Figure 13D). These data showed that the mean number of years separating identical core genomes was 2.5 years (range 0-15), and the mean temporal distances of identical genomes were similar within BC (2.9 years) and England (1.9 years) and between the two (2.7 years). The number of pairwise SNPs increased with temporal separation across all BC and English genomes from the same sublineage (Pearson’s Correlation 0.126, p<0.001), with a mean of 4.9 SNPs (range 0–23) separating genomes from the same year and sublineage, compared to 7.8 SNPs (range 6-11) after 19 years (Supplementary Figure 13D). This means that inference of direct patient-to-patient TPA transmission using the core genome will be challenging at the population level, and limit opportunities for real time genomic epidemiology because identical genomes can be separated by many years, and confidence intervals around temporal reconstructions will be broad. In the case of sublineage 14, we first detected this in BC, then the following year in England. Since we had a deeply sampled survey of populations over time for both countries, it seems likely that this represents a novel introduction into BC and England. However, low temporal rates and incomplete sampling, mean this must be interpreted with caution.

We also found some rarer sublineages – either as singleton strains, or those private to a single country. Whilst this might be expected in poorly sampled and geographically distant locations, such as Cuba, Haiti, Mexico and Zimbabwe, we found that the majority of private (6/8) and singleton (5/8) sublineages were actually from Canada, the UK or USA (Figure 4A), suggesting deeper sampling elsewhere will also find novel diversity.

Given our observations of individual sublineage expansion, we investigated whether the expansion could be related to antimicrobial resistance. Overlaying inferred macrolide resistance causing SNPs (A2058G, A2059G) in the ribosomal 23S rRNA gene on these population expansions showed evidence of macrolide resistance in 6/9 multi-country sublineages (Supplementary Figure 14), with the majority of samples being resistant in the largest sublineages 1, 2, 8 and 14. In contrast, only one private sublineage (sublineage 6, n=2) contained a macrolide resistant sample, suggesting that macrolide resistance is potentially linked to expansion in multicountry sublineages.

## Discussion

Previous attempts to understand the origins of the original syphilis pandemic^3,6,20^, as well as the dynamics of the current one^3,4^, have been constrained by the technical difficulty of sequencing TPA genomes, as well as relatively small datasets, with limited geographical diversity and sampling biases. In our study, we assembled the most comprehensive and broad ranging collection of syphilis samples from around the world to date, including samples from both the 20^th^ and 21^st^ century. Despite this, we still find that the TPA population consists of just two deep branching lineages, SS14-lineage and Nichols-lineage, with no outlying lineages. We were able to show that these lineages are both globally distributed and, where we have densely sampled, we find the relative proportions of each to be consistent. Although the overall diversity detected within the Nichols-lineage is far greater than that of SS14-lineage, suggesting earlier dissemination, we also found that these two major lineages exhibit similar phylodynamics, with recent sublineage expansions apparent in both lineages. This suggests that both Nichols- and SS14-lineages are capable of exploiting the transmission pathways driving the current syphilis epidemic.

Amongst our data, we sequenced the first genomes from syphilis patients in Africa, and our analysis shows that these genomes represented novel private sublineages, but their genomic diversity is nested entirely within the existing phylogenetic framework – these TPA genomes are not unusual. Indeed, we even observed the same pattern of Nichols- and SS14-lineages both being present in Zimbabwe, suggesting multiple introduction events into Zimbabwe. The same was true for genomes sequenced from Central Russia (Tuva Republic), where the private sublineage 5 represented novel, but entirely unremarkable genomic diversity.

In our study, we found that sublineages and closely related samples were more likely to be shared among deeply sampled countries. This suggests that sublineage sharing between countries is high, and deeper sampling of other countries will likely reveal similar patterns of sharing. As sampling depth increased we also found more rare sublineages, notably sublineage 6, representing novel outlying genetic diversity basal to all contemporary Nichols-lineage examples, in 2/240 contemporary UK patients. This suggests that some sublineages may truly be rare, whilst the high frequency of other sublineages could reflect either fitness advantages or epidemiological factors such as infecting patients within particularly high-risk sexual networks, allowing these sublineages to expand more successfully. Singleton or private sublineages could reflect insufficient sampling of a country or region, or sampling biases within a country (e.g. published genomes from Portugal^17^ came from a single clinic in Lisbon). These sublineages may therefore reflect transmission networks that are either contained within a less internationally mobile demographic, or may reflect transmission networks common in a region that is otherwise poorly sampled (e.g. Africa).

Our observation that the well-studied Nichols reference genomes (largely derived from or related to the original Nichols-1912 isolate) form an isolated clade, not represented in contemporary TPA, is important. One possible explanation is that these samples form a distinct clade due to convergent evolution in the rabbit model. However, this clade contains samples both extensively and minimally passaged, whilst other samples passaged in rabbits are distributed throughout the broader phylogeny, included in three SS14 sublineages (1, 2, 4) and two additional Nichols sublineages (7, 8). This indicates that passage in the rabbit model has not overly affected other parts of the phylogeny. The majority of Nichols-lineage strains collected prior to 1988 belong to this clade, and these samples mostly come from a small group of laboratories in the USA. Therefore, it is also tempting to suggest that this reflects a sampling bias for that time period. However, the phylogenetic placement of TPA_AUSMELT-1 within the same clade, isolated in 1977 in Australia^29^, and independently cultured and sequenced, contradicts this hypothesis, and may suggest that this clade represents the dominant TPA of the period. The complete absence of related genomes in contemporary sampling could represent a decline to becoming a rare or even extinct lineage and therefore implies that the Nichols reference strain is not representative of contemporary syphilis, or even contemporary Nichols-lineage strains.

Our data show that for some sublineages, modern syphilis is a truly global disease, with shared lineages, sublineages and indeed nearly identical strains all over the world. The large expansions of highly related genomes, in particular sublineage 1, represent the bulk of sequenced genomes in our dataset, and the widespread sharing of major lineages suggests we have sampled from a series of globally contiguous sexual networks, making contemporary syphilis effectively panmictic.

Furthermore, we find evidence of a striking change in the genetic diversity and effective population size of TPA genomes, suggestive of a possible population bottleneck occurring between the late 1990s and early 2000s. This was followed by a rapid expansion of certain sublineages, leading to the contemporary TPA population structure. This bottleneck, potentially a consequence of post-HIV safe sex messaging, persistent antimicrobial prophylaxis in at risk HIV positive populations, and possibly HIV-associated mortality, appears to have led to a striking duality in the TPA dominating populations before and after. The rapid expansion may be attributed to a relaxation of sexual behaviour following the widespread introduction of highly active antiretroviral therapy. Notably, although macrolide resistance was not universally distributed throughout the phylogeny or present in all sublineages, most of the multi-country sublineages were largely macrolide resistant, and this could also have played a role through off-target effects, e.g. during treatment of other (particularly sexually transmitted) infections^31^. Azithromycin and other macrolides are no longer recommended treatments at any stage of syphilis in the European syphilis management guidelines^32^.

There have been some documented reports of clinically diagnosed syphilis caused by *Treponema pallidum* subspecies *endemicum* (TEN)^33,34^. In our study, most novel genomes were clinically diagnosed and confirmed by diagnostic PCRs that do not discriminate between subspecies, yet we found only TPA. Therefore, although we cannot rule out that TEN causes syphilis due to the limits of our sampling framework, our data suggest TEN is not a major contributor to the burden of syphilis in any of our well sampled countries.

Our study has a number of limitations. Our samples were collected in an opportunistic manner, using residual samples available in regional or national archives. Since our sampling coverage is uneven, with some countries either missing or under-sampled, it was not possible to infer the direction of transmission between countries. Nevertheless, we provide a snapshot of strains from Africa, Asia and South America, all of which overlap with the genetic diversity of our more deeply sampled regions (North America and Europe), suggesting we have captured key global lineages. We also show that even deeply sampled countries can harbour rare sublineages, and it is therefore likely that future studies will reveal further novel diversity. We were constrained in our ability to obtain and sequence older samples from prior to the widespread adoption of molecular diagnostics in the early 2000s, and this is largely influenced by the difficulty in isolating new strains prior to the recent development of *in vitro* culture^21^, and the lack of, and long-term storage of clinical swabs. Most (but not all) older strains come from the USA, and this could mean that we do not accurately reflect the global population structure prior to 2000.

Despite this our data show the *T. pallidum* infecting patients today is not the same *T. pallidum* infecting patients even 30 years ago – ancestral sublineages appear to have become extinct, being replaced by new sublineages that have swept to dominance across the globe with the dramatic upswing in syphilis cases in the US, UK, and other Western European countries, which were heavily sampled in our study. That such a bottleneck is linked to HIV-related behavioural change during the 1990s, rather than the introduction of antibiotics after the Second World War, further supports the importance of sexual behaviour in transmission dynamics. In future work, it would be interesting to integrate epidemiological evidence of sexual networks in purpose designed cohort studies to explore this further.

## Methods

### Samples

Ethical approval for all clinical samples was granted by the London School of Hygiene and Tropical Medicine Observational Research Ethics Committee (REF#16014) and the National Health Service (UK) Health Research Authority and Health and Care Research Wales (UK; 19/HRA/0112).

Novel samples from Australia (Brisbane, Melbourne), Belgium (Antwerp), Canada (Alberta, British Columbia), Hungary (National collection), Ireland (Dublin), Russia (Tuva Republic), South Africa (Johannesburg), Spain (Barcelona), Sweden (National collection), the UK (National collection), Zimbabwe (3 regions), were sequenced directly from genomic DNA extracted from clinical patient swabs or biopsies, typically utilising de-identified residual diagnostic samples which were further pseudonymised before analysis. Additional novel samples from Australia (Melbourne), Haiti and the USA (6 cities) were sequenced from historic freezer archives after prior passage in the rabbit model^4^.

DNA extracts were quantified by qPCR (TPANIC_0574) as previously^4^, and grouped into pools of either 32 or 48 with similar (within 2 CT) treponemal load with high concentration outlier samples diluted as necessary. We added 4μl pooled commercial human gDNA (Promega) to all samples to ensure total gDNA > 1μg/35μl, sufficient for library prep.

### Sequencing

Extracted genomic DNA was sheared to 100-400 bp (mean distribution 150 bp) using an LE220 ultrasonicator (Covaris Inc). Libraries were prepared (NEBNext Ultra II DNA Library prep Kit, New England Biolabs, Massachusetts, USA) using initial adaptor ligation and barcoding with unique dual indexed barcodes (Integrated DNA Technologies, Iowa, USA). Dual indexed samples were amplified (6 cycles of PCR, KAPA HiFi kit, Roche, Basel, Switzerland), quantified (Accuclear dsDNA Quantitation Solution, Biotium, California, USA), then pooled in preassigned groups of 48 or 32 to generate equimolar pools based on Total DNA concentration. 500 ng pooled DNA was hybridised using 120-mer RNA baits (SureSelect Target enrichment system, Agilent technologies; Bait design ELID ID 0616571)^4,35^. Enriched libraries were sequenced on Illumina HiSeq 4000 to generate 150 bp paired end reads at the Wellcome Sanger Institute (Cambridgeshire, UK) as previously described^36^. For one rabbit passaged sample from Melbourne, Australia (TPA_AUSMELT-1)^29^, genomic DNA extracted from historically archived tissue lysate was sequenced on Illumina NextSeq 500 (150 bp paired end reads, Nextera DNA Flex libraries) without any prior enrichment to an estimated 1Gb/sample at the Doherty Institute (Melbourne, Australia).

### Sequence analysis and Phylogenetics

We filtered *Treponema* genus-specific sequencing reads using the full bacterial and human Kraken 2^37^ v2.0.8 database (March 2019), followed by trimming with Trimmomatic^38^ v0.39 and downsampling to a maximum of 2,500,000 using seqtk v1.0 (available at https://github.com/lh3/seqtk) as previously described^4^. For publicly available genomes, raw sequencing reads were downloaded from the European Nucleotide Archive (ENA) and subjected to the same binning and downsampling procedure. For five public genomes (see Supplementary Data 1), raw sequencing reads were not available; for these we simulated 150 bp PE perfect reads from the RefSeq closed genomes using Fastaq (available at https://github.com/sanger-pathogens/Fastaq).

For phylogenomic analysis, we mapped *Treponema*-specific reads to a custom version of the SS14_v2 reference genome (NC_021508.1), after first masking 12 repetitive Tpr genes (Tpr A-L), two highly repetitive genes (arp, TPANIC_0470), and five FadL homologs (TPANIC_0548, TPANIC_0856, TPANIC_0858, TPANIC_0859, TPANIC_0865) using bedtools^39^ v2.29 maskfasta (positions listed in Supplementary Data 2). We mapped prefiltered sequencing reads to the reference using BWA mem^40^ v0.7.17 (MapQ ≥ 20, excluding reads with secondary mappings) followed by indel realignment using GATK v3.7 IndelRealigner, deduplication with Picard MarkDuplicates v1.126 (available at http://broadinstitute.github.io/picard/), and variant calling and consensus pseudosequence generation using samtools^41^ v1.6 and bcftools v1.6, requiring a minimum of two supporting reads per strand and five in total to call a variant, and a variant frequency/mapping quality cut-off of 0.8. Sites not meeting our filtering criteria were masked to ‘N’ in the final pseudosequence. After mapping and pseudosequence generation, we repeated the masking of the 19 genes on the final multiple sequence alignment using remove_block_from_aln.py available at https://github.com/sanger-pathogens/remove_blocks_from_aln/ to ensure sites originally masked in the reference were not inadvertently called with SNPs due to mapped reads overlapping the masked region. These 19 regions of recombination and genomic uncertainty due to gene orthology or repetitive regions^3,4,6^ accounted for 30,071 genomic sites (Supplementary Data 2).

For basic lineage assignment of genomes, we excluded sequences with >75% genomic sites masked to ‘N’. A SNP-only alignment was generated using snp-sites^42^, and a maximum likelihood phylogeny was calculated on the variable sites using IQ-Tree^27^ v1.6.10, inputting missing constant sites using the ‘-fconst’ argument, and using a general time reversible (GTR) substitution model with a FreeRate model of heterogeneity^43^ and 1000 UltraFast Bootstraps^44^.

For finescale analysis of high-quality genomes, we excluded sequences with >25% genomic sites masked to ‘N’ (i.e. >75% genomic sites passing filters at >5x and not masked). We used Gubbins^26^ v2.4.1 (20 iterations) to generate recombination-masked full genome length and SNP-only alignments. Gubbins^26^ identified 19 further putative regions of recombination affecting 2.1% of genomic sites (n=23,567) and 27 genes (listed in Supplementary Data 2), meaning we masked a maximum of 4.7% (53,638 sites) of the genome over the 38 regions. We used IQ-Tree on the SNP-only alignment containing 901 variable sites, inputting missing constant sites using the ‘-fconst’ argument, and allowing the built-in model test to infer a K3Pu+F+I model and 10,000 UltraFast bootstraps.

To cluster genomes, we initially performed joint ancestral reconstruction^45^ of SNPs on the phylogeny using pyjar (available at https://github.com/simonrharris/pyjar), and used this to determine phylogenetic clusters with a 10 SNP threshold in rPinecone^28^ (available at https://github.com/alexwailan/rpinecone). We further investigated this by using IQ-tree to generate 100 standard non-parametric bootstraps on the maximum likelihood phylogeny, and used the resulting 100 trees as independent inputs to rPinecone, as described in the rPinecone manuscript^28^. We used the hierarchical clustering ‘hclust’ algorithm in R^46^ to group rPinecone clusters, and evaluated different proportions of trees supporting clusters against the phylogeny (Supplementary Figure 3).

For temporal analysis, our dataset was too large for robust model testing of all genomes, so we stratified our dataset by sublineage and country, then used the random sampler in R^46^ to select a maximum of five genomes from each strata, plus all singleton strains, yielding a dataset of 138. We extracted the sequences from the multiple sequence alignment using seqtk and the subtree from our broader phylogeny using ape^2247^ v 5.4.1 ‘keep.tip’. Root-to-tip distance analysis from this subtree showed a correlation of 0.327 and R^2^ of 0.11 (Supplementary Figure 15), and we proceeded to BEAST analysis. We initially ran BEAST^30,48^ v1.8.4 on our recombination-masked SNP-only alignment containing 592 variable sites, correcting for invariant sites using the constantPatterns argument, in triplicate using both a Strict Clock model^49^ (starting rate prior 1.78E^-7^) and an Uncorrelated Relaxed Clock model^50^, with HKY substitution model^51^ and diffuse gamma distribution prior^52^ (shape 0.001, scale 1000) over 100 million MCMC cycles with 10 million cycle burnin. We evaluated constant, relaxed lognormal, exponential and Bayesian Skyline (10 categories) population distributions^53^. All MCMC chains converged with high effective sample sizes, and on inspection of the marginal distribution of ucld.stdev we could not reject a Strick Clock. We used the marginal likelihood estimates from the triplicate BEAST runs as input to Path Sampling and Stepping Stone Sampling analysis^54,55^, and this suggested that the Strict Clock with Bayesian Skyline was the optimal model for this dataset (Supplementary Figure 16), with an inferred molecular clock rate of 1.23 ×10^−7^ substitutions/site/year. To ensure that our findings were not artefactual to the down-sampled dataset, we re-stratified the dataset by sublineage, country and year, selecting a maximum of 3 genomes from each stratum, plus all singleton strains, yielding a dataset of 168 genomes with 466 variable sites. We ran BEAST on this new subsampled dataset using the optimal Strict Clock (with starting rate prior of 1.23 ×10^−7^, inferred from the previous analysis) with Bayesian Skyline from above, with the equivalent results (Supplementary Figure 17).

To evaluate the temporal dynamics of sublineages, we tested the temporal signal for the 4 largest sublineages 1, 2, 8, and 14 (Supplementary Figure 10). Sublineage 14 had poor temporal signal and was excluded from further analysis. We performed independent BEAST analyses on the remaining sublineage multiple sequence alignments using the optimal Strict Clock model with Bayesian Skyline (10 population groups), evaluating 3 independent chains of 200 million cycles for each.

To analyse the full dataset (n=520 after excluding heavily passaged samples or those with uncertain collection dates, 883 variable sites), after evaluating temporal signal (Supplementary Figure 18), we initially attempted to reproduce our model in BEAST 1.8.4, but this proved unachievable with our local implementation and compute arrangements. To analyse the full dataset, we therefore reconstructed the optimal BEAST v1.8.4 model (Strict Clock with reference rate prior of 1.23 ×10^−7^s/s/y, Coalescent Bayesian Skyline distribution with 10 populations^51,53^) in a BEAST2^56^ v2.6.3 implementation with BEAGLE^48^ libraries optimised for Graphical Processing Units, analysing the 520 genomes over 500 million MCMC cycles in triplicate.

To further confirm the temporal signal in our full 520 genome tree, we used the TIPDATINGBEAST^57^ package in R to perform a date randomisation test, generating 20 new datasets with randomly reassigned dates – BEAST2 analysis using the same prior conditions found no evidence of temporal signal in these replicates, indicating that the signal in our tree was not found by chance (Supplementary Figure 19).

We used logcombiner to generate consensus log and treefiles, and treeannotator to create median maximum credibility trees. We generated Bayesian Skyline and Lineage accumulation plots using the combined log and tree files in Tracer v1.7.1^30^, exporting the data for subsequent plotting in R. To evaluate the posterior distribution of population expansion times, we used the script population_increase_distribution_BEAST.py (available at https://github.com/chrisruis/tree_scripts), which uses the BEAST log and tree files to identify the first increase in relative genetic diversity from the PopSizes columns and the date of this increase using the corresponding number of nodes in the GroupSizes columns and the node heights in the respective tree. We required a 2-fold population expansion (defined by setting ‘-p’ to 100). The script outputs the proportion of trees in the posterior distribution that support an increase in relative genetic diversity, along with the distribution of expansion dates, which we plotted in R^46^. We repeated this analysis using the script population_change_support_BEAST.py (available at https://github.com/chrisruis/tree_scripts), which looks for an increase or decrease of effective population size within a defined window, testing for supported start dates of a 2-fold population decline or expansion between 1990 and 2015.

Macrolide resistance alleles were inferred using the competitive mapping approach previously described^4,36^ (available at https://github.com/matbeale/Lihir_Treponema_2020). To infer pairwise SNP distances between samples, we used pairsnp (available at https://github.com/gtonkinhill/pairsnp). We constructed networks of minimum pairwise distance and shared lineages in R^46^, and plotted these as heatmaps using ggplot2^58^. Nucleotide diversity (π) for different clades was inferred from the multiple sequence alignments using EggLib^59^ v3.0.0b21, including variable sites present in at least 5% of genomes. For geospatial analysis, we used the CoordinateCleaner^60^ v2.0-17 package in R to define the centroid position for each country, apart for Russia (where we used the centroid of the Tuva Republic) and Mexico (where we used Mexico City). Geographic distances between countries (using the country centroid or location defined above) were determined using the distVincentyEllipsoid function from the geosphere^61^ v1.5-10 package. Correlations between pairwise genetic, geographic and temporal distance were inferred using Pearson’s R Correlation via the ‘cor’ function in R, where we compared ‘real’ correlation with 1000 bootstraps resampled with replacement to obtain a p value. Sample counts were plotted using ggmaps^62^ over maptiles downloaded from Stamen Design (http://maps.stamen.com). All phylogenetic trees were plotted in R using ggtree^63^. All figures used ggplot2^58^ v3.3.2 for plotting.

## Data Availability

Sequencing reads for all novel genomes have been deposited at the European Nucleotide Archive in BioProjects PRJEB28546, PRJEB33181 and PRJNA701499. All accessions, corresponding pseudonymised sample identifiers and related metadata are available in Supplementary Data 1.

https://www.ebi.ac.uk/ena/browser/view/PRJEB28546

https://www.ebi.ac.uk/ena/browser/view/PRJNA701499

## Data Availability

Sequencing reads for all novel genomes have been deposited at the European Nucleotide Archive in BioProjects PRJEB28546, PRJEB33181 and PRJNA701499. All accessions, corresponding sample identifiers and related metadata are available in Supplementary Data 1.

## Acknowledgements

The authors acknowledge the sequencing team at the Wellcome Sanger Institute, and Christoph Puethe and the Pathogen Informatics team for computational support. We thank additional technical staff involved in sample diagnostics, DNA extraction and sample retrieval in laboratories at Public Health England and NHS laboratories, UK; British Colombia CDC and Alberta Precision Laboratories, Canada; National Public Health Center, Budapest, Hungary; FRC Kazan Scientific Center, Tuva, Russia; National Institute for Communicable Diseases, Johannesburg, South Africa; Institute of Tropical Medicine, Antwerp, Belgium; Sahlgrenska University Hospital, Gothenburg, Sweden; Hospital Vall d’Hebron, Barcelona, Spain; Midlands Regional Hospital Portlaoise, Ireland; Pathology Queensland Central Laboratory, Australia; WHO Collaborating Centre for Gonorrhoea and other STIs, Sweden. We thank G. Tonkin-Hill, A. van Tonder, and members of the Thomson team for helpful discussions during analysis. MAB and NRT are supported by Wellcome funding to the Sanger Institute (#206194). MM is funded by the UKRI and NIHR [COV0335; MR/V027956/1, NIHR200125] and the EDCTP [RIA2018D-249]. DMW is funded by a Queensland Advancing Clinical Research Fellowship from the Queensland Government. DAW is supported by an Investigator Grant (1174555) from the National Health and Medical Research Council of Australia. SAL is funded by the National Institutes of Health (R01 AI42143). This research was funded in whole, or in part, by the Wellcome Trust (#206194). For the purpose of Open Access, the author has applied a CC-BY public copyright licence to any Author Accepted Manuscript version arising from this submission.

## Author Contributions

Conceived and designed the study: MAB, MM, SAL, NRT. Coordinated collaboration, receipt and sequencing of samples: MM, MAB, MU. Collected, retrieved and prepared samples and patient metadata: MM, MJC, M-KL, RP, EB, TC, ME, CFN, AG, MG, CRK, RKh, RKu, MA, BJM, AO, EP, FP, CRi, DR, SS, ESm, ELS, GT, JHV, CW, DMW, DAW, GH, PN, MK, MU, SAL, MGM, HF. Performed laboratory work for sequencing: MAB, GT. Analysed the data: MAB. Provided analytical tools and advice: CRu. Wrote the initial draft of the paper: MAB, with substantial contributions from NRT. All authors viewed and contributed to the final paper.

## Supplementary Figures

**Supplementary Figure 1. Phylogenetic distribution of 726 *Treponema pallidum* ssp *pallidum* partial genomes**. Maximum likelihood phylogeny of 726 partial (>25% of genome positions) genomes shows two primary lineages (Nichols, SS14), with no obvious correlation by country or continent. Tree tip points are coloured by continent, while coloured strips show continent, country and TPA lineage. One very low coverage sample (TPA_BCC144, Canada, 47% genome breadth, 7.9X mean coverage) appears basal to the SS14-lineage clade in this phylogeny, but due to low coverage it was not possible to determine the correct placement.

**Supplementary Figure 2. Finescale analysis of 528 high quality (>75% reference sites) TPA genomes and sublineages**. Recombination masked WGS phylogeny of 528 genomes. Tree tips and coloured strips show sublineage.

**Supplementary Figure 3. Evaluating phylogenomic clustering using bootstrap resampled trees**. We generated 100 bootstraps from our finescale analysis of 528 TPA genomes, independently running rPinecone (10 SNP threshold) on each bootstrapped tree. Hierarchical clustering was used to group rPinecone sublineages, and we applied different support thresholds (minimum % of trees remaining) to explore the consistency of sublineages. Nichols-sublineages were all well supported, but some SS14-sublineages lacked support in many bootstraps. To focus on the more stable sublineages we required that at least 5% of the bootstrap replicates supported a cluster. Plot shows maximum likelihood phylogeny, with metadata columns showing cluster assignment along the x-axis for the original maximum likelihood cluster assignment, then allowing for 95%, 80%, 50%, 20% and 5% of bootstrap variation observed. Final sub-lineage assignments are shown against the 5% cluster assignments. Note that non-zero branch lengths were added by IQ-Tree during maximum likelihood tree estimation, leading to an artifactual ladder-like appearance for sublineage 1.

**Supplementary Figure 4. Detailed subtree of SS14-lineage**. Recombination masked WGS phylogeny, showing the SS14-lineage and sublineages. The low diversity globally distributed sublineage 1 has been collapsed to enable visualization of the remaining sublineages. Tip points are coloured by sublineage, and coloured strips show sublineage and country. Blue triangle indicates collapsed Nichols-lineage, pink triangle indicates collapsed sublineage 1. Two samples close to the root of the common SS14-lineage clades were clustered as sublineage 1, and are shown.

**Supplementary Figure 5. Subtree highlighting novel Nichols-lineage strains**. Recombination masked WGS phylogeny, showing the Nichols-lineage and sublineages. Tip points are coloured by sublineage, and coloured strips show sublineage and country. Shaded boxes highlight basal Nichols-lineage outgroup sublineages 6 and 7. The large clonal sublineage 14 has been collapsed to enable clearer visualization of the remaining taxa. The pink triangle indicates collapsed SS14-lineage, blue triangle indicates the collapsed sublineage 14.

**Supplementary Figure 6. Commonly used Nichols Reference genomes form a monophyletic clade unrelated to contemporary clinical strains**. A-Recombination masked WGS phylogeny, showing the Nichols-lineage and sublineages. Shaded grey box shows a monophyletic clade containing commonly used reference genomes as well as genetically related strains. Tip points are coloured by sublineage, and coloured strips show sublineage and country. Pink triangle indicates collapsed SS14-lineage. B-Expanded view of a seemingly extinct clade containing common reference strains including Nichols_v2, DAL-1 and Seattle_Nichols. The most recent sample closely related to the reference strains (TPA_USL-SEA-83-1) was collected in 1983, while the latest sample for the entire clade (TPA_USL-Phil-3) was collected in 1987. The provenance of the sample originally used for sequencing the DAL-1 genome is uncertain, but in the literature the original isolation was in 1988. The placement of both DAL-1 and TPA_USL-SEA-83-1 within the diversity of Nichols-1912 derivatives suggests the possibility of the samples being mislabeled in the handling laboratories.

**Supplementary Figure 7. Finescale analysis of 528 high quality TPA genomes and sublineages, showing distribution of samples sequenced directly from clinical samples and those passaged in rabbit model**. A – Whole genome phylogeny showing distribution of samples sequenced directly from clinical sample or rabbit-passaged. B – Distribution of samples sequenced directly from clinical sample and rabbit-passaged samples according to sublineage. Samples passaged in rabbits are distributed throughout the global TPA phylogeny, and present in 9/17 sublineages. Older samples from before 2000 were isolated via rabbit passage, and dominate extinct clusters, as well as clustering close to the most recent common ancestor of contemporary sublineages such as SS14 sublineage 1.

**Supplementary Figure 8. Bayesian Skyline analysis of population decline and expansion start dates**. Plots show posterior distribution of supporting trees for the start of either a 2-fold decline (pink) or expansion (blue) using a scanning approach within a window of 1990-2015. Analysis provides strong support for a population bottleneck in or around 2000, and moderate support for a subsequent expansion after 2010. Population changes are scaled to the population size averaged over the starting period for each tree. Therefore, if a particular tree already exhibited a decline near the starting timepoint, this may mean this tree does not show expansion, resulting in reduced overall support for expansion.

**Supplementary Figure 9. Bayesian Skyline analysis of sublineages**. Plots show population expansions occurring during the early 2000s for all sublineages with >15 samples apart from sublineage 14. Sublineage 14, which had low temporal signal, did not converge after multiple BEAST runs. Shows Skyline plots of sublineages 1, 2, 8 and plot for all samples from Figure 5.

**Supplementary Figure 10. Subtrees of major sublineages, with corresponding root-to-tip distance plots**. All subtrees showed some evidence of temporal signal, but this was very weak for the recently emerged sublineage 14. Graphs are annotated with slope and time to most recent common ancestor (TMRCA) inferred directly from the maximum likelihood subtree, not BEAST.

**Supplementary Figure 11. Finescale analysis of 528 high quality TPA genomes and sublineages, highlighting private and singleton sublineages**. Private and singleton sublineages are nested within the existing diversity of the TPA phylogeny. Tip points indicate sublineage, coloured tracks highlight singletons or private sublineages (with corresponding sublineage number), and country.

**Supplementary Figure 12. Effect of geographic distance on genetic distance**. A-Pairwise comparison of genetic distance (SNPs) and geographic distance (km; calculated using country centroids) within Nichols- and SS14-lineages, including linear regression (95% CI not visible). B-Pairwise comparison of genetic distance (SNPs) and geographic distance (km; calculated using country centroids) within the four major multi-country sublineages (SS14: 1, 2; Nichols: 8, 14). Includes linear regression (95% CI not visible)

**Supplementary Figure 13. Sharing of sublineages and closely related strains within and between British Columbia (Canada) and England (UK)**. A-Syphilis incidence per 100,000 population by year for British Columbia, (Canada) and England (UK) using currently published data. B-TPA sublineage counts for each year, using high quality genomes from British Columbia (n=84) and England (n=240). British Columbia samples collected from 2000-2018, English samples collected from 2012-2018. C-Pairwise comparison of SNP distance distributions from samples within and between British Columbia and England. D-Comparison of SNP distance and temporal distance within and between British Columbia and England. The plot is divided into hexagonal bins, with the colour of each hexagon representing the number of comparisons (white=none, purple=few, green=many, see scale). Linear regression lines also shown (95% CI not visible).

**Supplementary Figure 14. Multicountry sublineages are broadly macrolide resistant**. A-Whole genome phylogeny showing distribution of macrolide resistance conferring SNPs (A2058G and A2059G). B-Distribution of macrolide resistance SNPs by sublineage, indicating number of samples per sublineage, and sublineage type. Note that while the common A2058G mutation was found in six sublineages (both Nichols- and SS14-lineages), we also found the less common A2059G in both SS14-lineage (sublineages 1, 2) and Nichols-lineage (sublineage 6).

**Supplementary Figure 15. Maximum Likelihood phylogeny of 138 representatively subsampled genomes**. A-Maximum likelihood phylogeny of 138 genomes randomly sampled to be representative of sublineage and country. B-Scatterplot showing root-to-tip distance against collection date, illustrating temporal signal in the dataset. C-Expanded version of B, showing regressed x-intercept.

**Supplementary Figure 16. Bayesian maximum credibility phylogeny of 138 representative genomes shows population contraction during the 1990s, followed by rapid expansion from the early 2000s onwards**. A-Time-scaled phylogeny of 138 genomes randomly sampled to be representative of sublineage, country and collection year. Coloured tracks indicate sublineage, country and collection year. Node points are shaded according to posterior support (black ≥96%, dark grey >91%, light grey >80%). Red bars on nodes indicate 95% Highest Posterior Density intervals. Blue line and shaded area highlights post-2000 expansion of lineages. B-Bayesian Skyline plot shows decline of effective population size after the second world war, flattening in the 1960s, followed by a sharp decline and rapid reemergence during the 1990s and 2000s.

**Supplementary Figure 17. Secondary BEAST analysis of 168 separately subsampled representative genomes**. A-Time-scaled phylogeny of 168 genomes randomly sampled to be representative of sublineage, country and collection year. Node points are shaded according to posterior support (black ≥96%, dark grey >91%, light grey >80%). Red bars on nodes indicate 95% Highest Posterior Density intervals. Blue line and shaded area highlights post-2000 expansion of lineages. B-Bayesian Skyline plot shows decline of effective population size after the second world war, flattening in the 1960s, followed by a sharp decline and reemergence during the 1990s and 2000s, indicative of a sharp bottleneck.

**Supplementary Figure 18. Maximum likelihood tree of 520 TPA genomes with minimal passage and robust collection dates, with corresponding root-to-tip distance plot**. Within the full tree, the temporal signal was weaker than in our subsampled dataset, but still plausible, given our prior analyses. Graphs are annotated with slope and time to most recent common ancestor (TMRCA) inferred directly from the maximum likelihood subtree, not BEAST.

**Supplementary Figure 19. Date Randomisation Test for full BEAST2 dataset confirms the temporal signal in the true dataset compared to 20 resampled datasets with randomly reassigned tipdates**. The median clock rate for the real dataset was 1.27 ×10^−7^, while all randomly assigned datasets gave substantially lower clock rates; the highest median clock rate for the randomized datasets was 8.02 ×10^−9^. Real sample (blue), randomized samples (pink).

**Supplementary Data 1**. Metadata and read accessions for all samples included in this study

**Supplementary Data 2**. Genomic regions masked due through prefiltering or recombination analysis.

