## Supplementary Figures for "Contemporary syphilis is characterised by rapid global spread of pandemic *Treponema pallidum* lineages"

**Beale *et al*, 2021**

**Supplementary Figures**

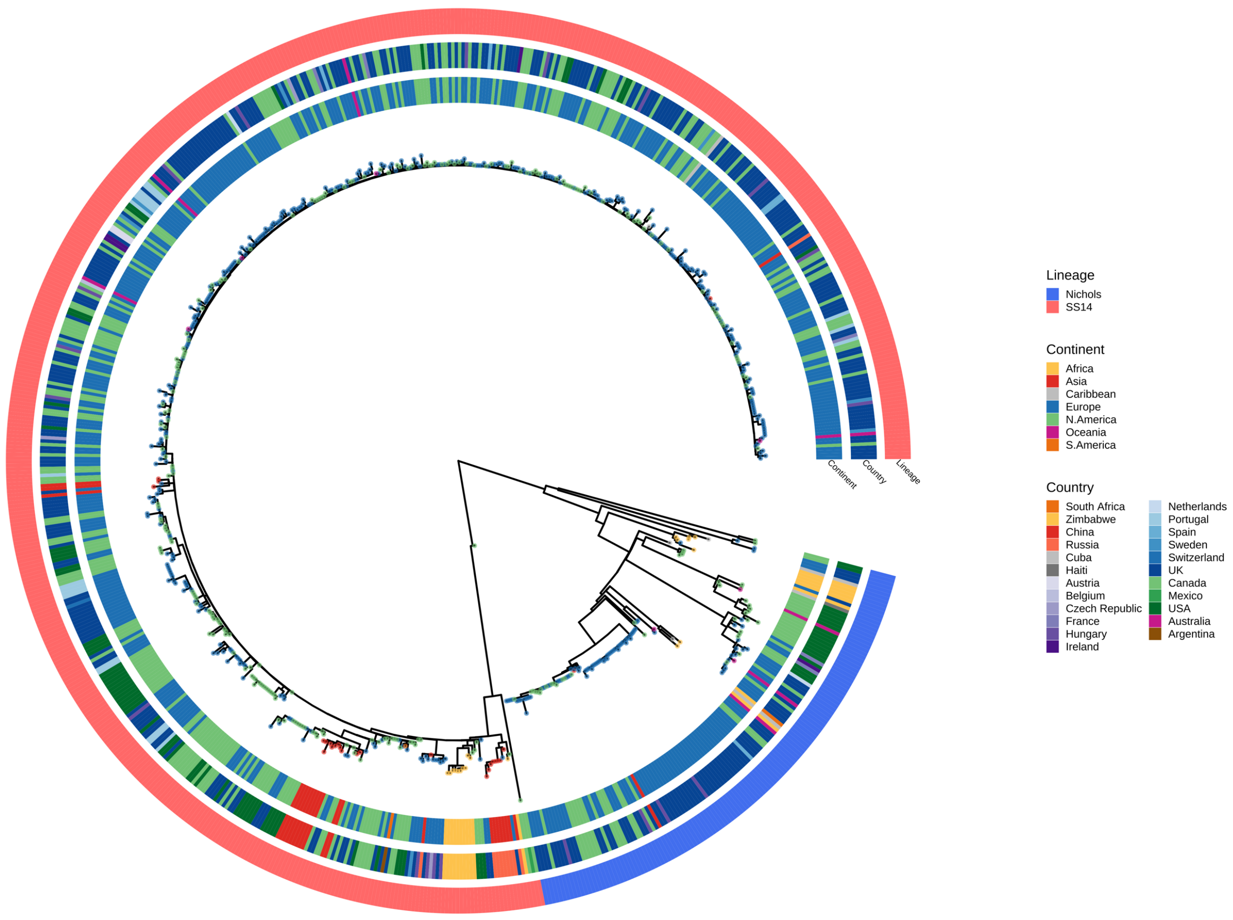


**Supplementary Figure 1. Phylogenetic distribution of 726 *Treponema pallidum* ssp *pallidum* partial genomes.** Maximum likelihood phylogeny of 726 partial (>25% of genome positions) genomes shows two primary lineages (Nichols, SS14), with no obvious correlation by country or continent. Tree tip points are coloured by continent, while coloured strips show continent, country and TPA lineage. One very low coverage sample (TPA_BCC144, Canada, 47% genome breadth, 7.9X mean coverage) appears basal to the SS14-lineage clade in this phylogeny, but due to low coverage it was not possible to determine the correct placement.

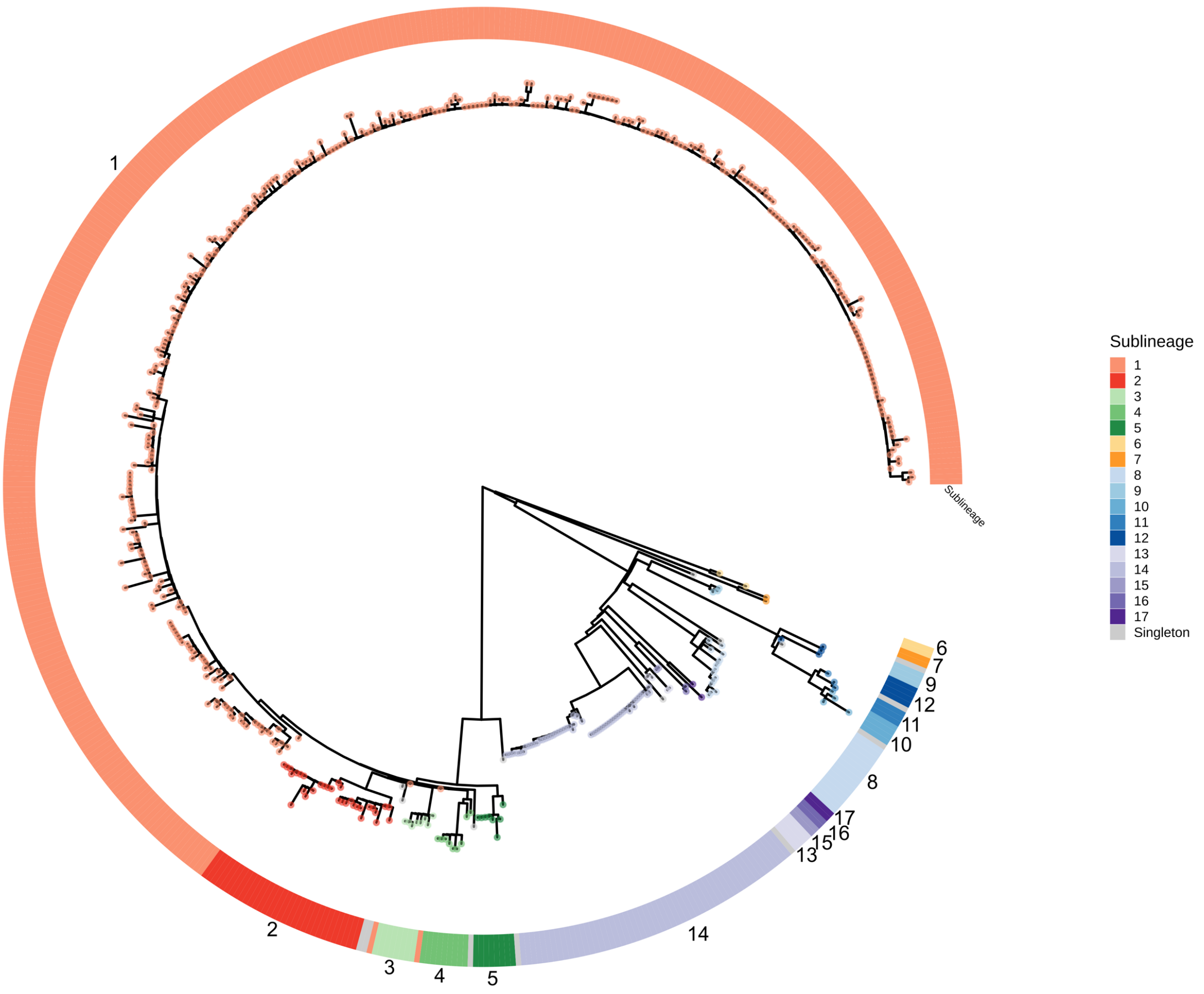


**Supplementary Figure 2. Finescale analysis of 528 high quality (>75% reference sites) TPA genomes and sublineages.** Recombination masked WGS phylogeny of 528 genomes. Tree tips and coloured strips show sublineage.

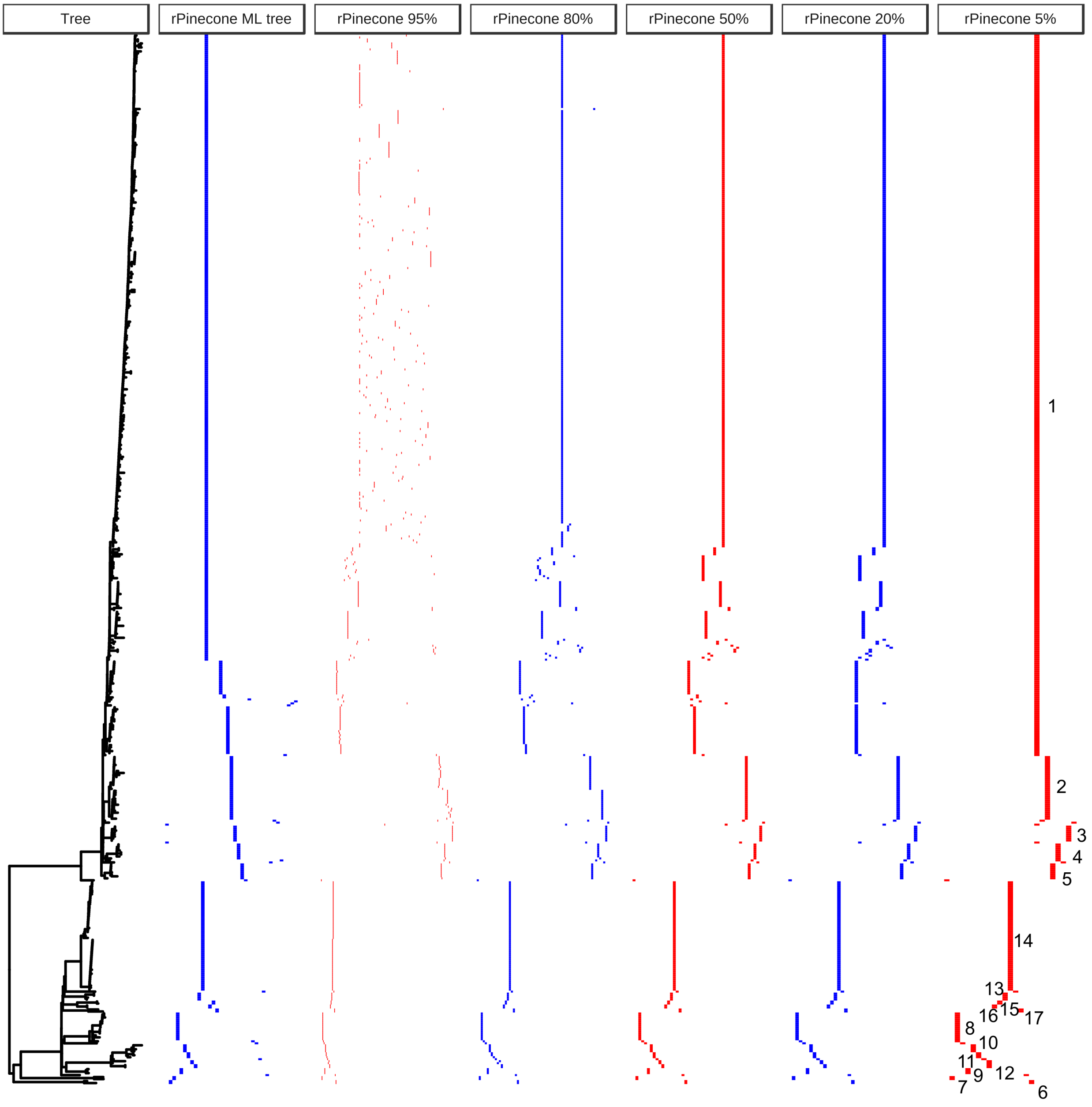


**Supplementary Figure 3. Evaluating phylogenomic clustering using bootstrap resampled trees.** We generated 100 bootstraps from our finescale analysis of 528 TPA genomes, independently running rPinecone (10 SNP threshold) on each bootstrapped tree. Hierarchical clustering was used to group rPinecone sublineages, and we applied different support thresholds (minimum % of trees remaining) to explore the consistency of sublineages. Nichols-sublineages were all well supported, but some SS14-sublineages lacked support in many bootstraps. To focus on the more stable sublineages we required that at least 5% of the bootstrap replicates supported a cluster. Plot shows maximum likelihood phylogeny, with metadata columns showing cluster assignment along the x-axis for the original maximum likelihood cluster assignment, then allowing for 95%, 80%, 50%, 20% and 5% of bootstrap variation observed. Final sub-lineage assignments are shown against the 5% cluster assignments. Note that non-zero branch lengths were added by IQ-Tree during maximum likelihood tree estimation, leading to an artifactual ladder-like appearance for sublineage 1.

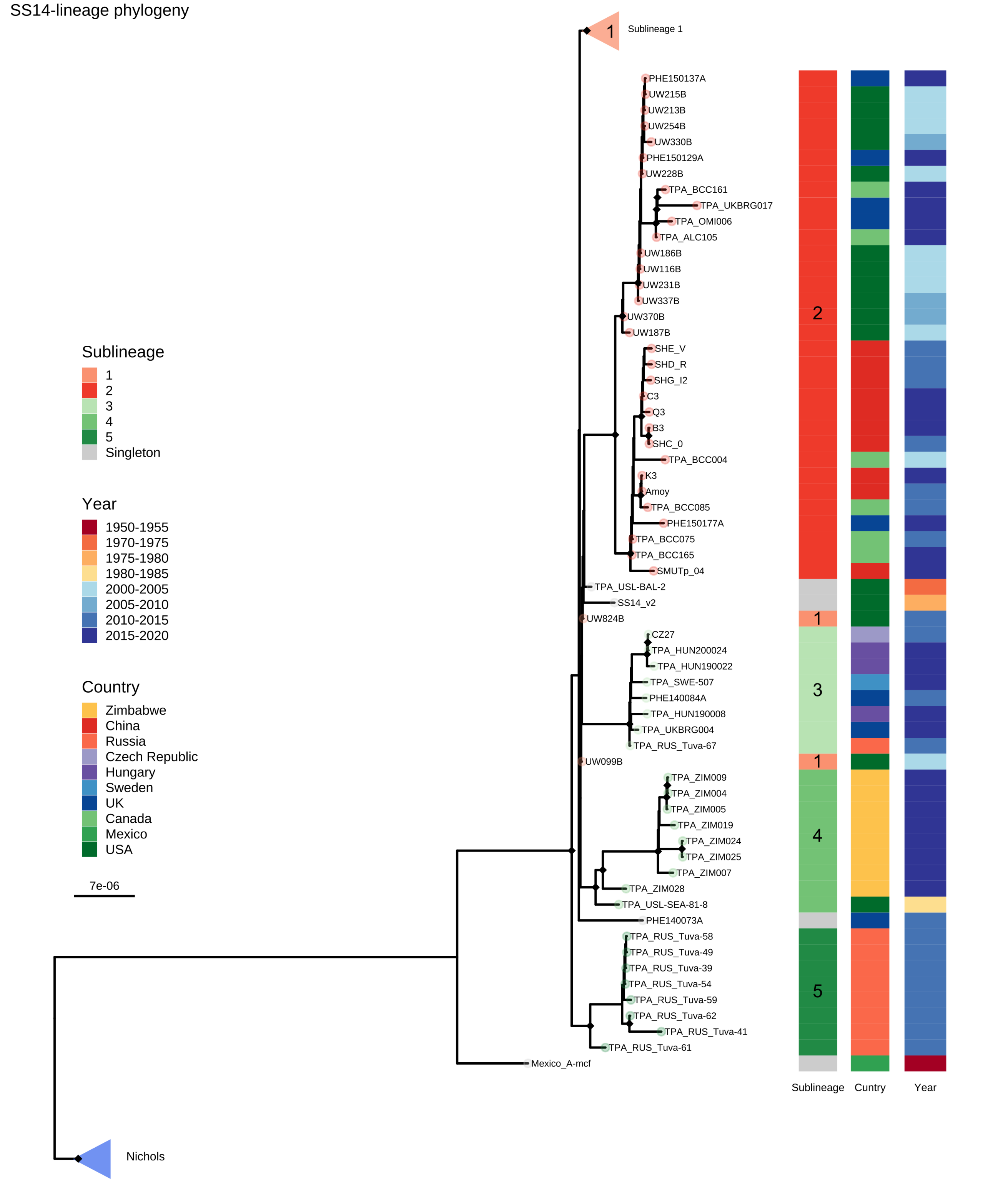


**Supplementary Figure 4. Detailed subtree of SS14-lineage.** Recombination masked WGS phylogeny, showing the SS14-lineage and sublineages. The low diversity globally distributed sublineage 1 has been collapsed to enable visualization of the remaining sublineages. Tip points are coloured by sublineage, and coloured strips show sublineage and country. Blue triangle indicates collapsed Nichols-lineage, pink triangle indicates collapsed sublineage 1. Two samples close to the root of the common SS14-lineage clades were clustered as sublineage 1, and are shown.

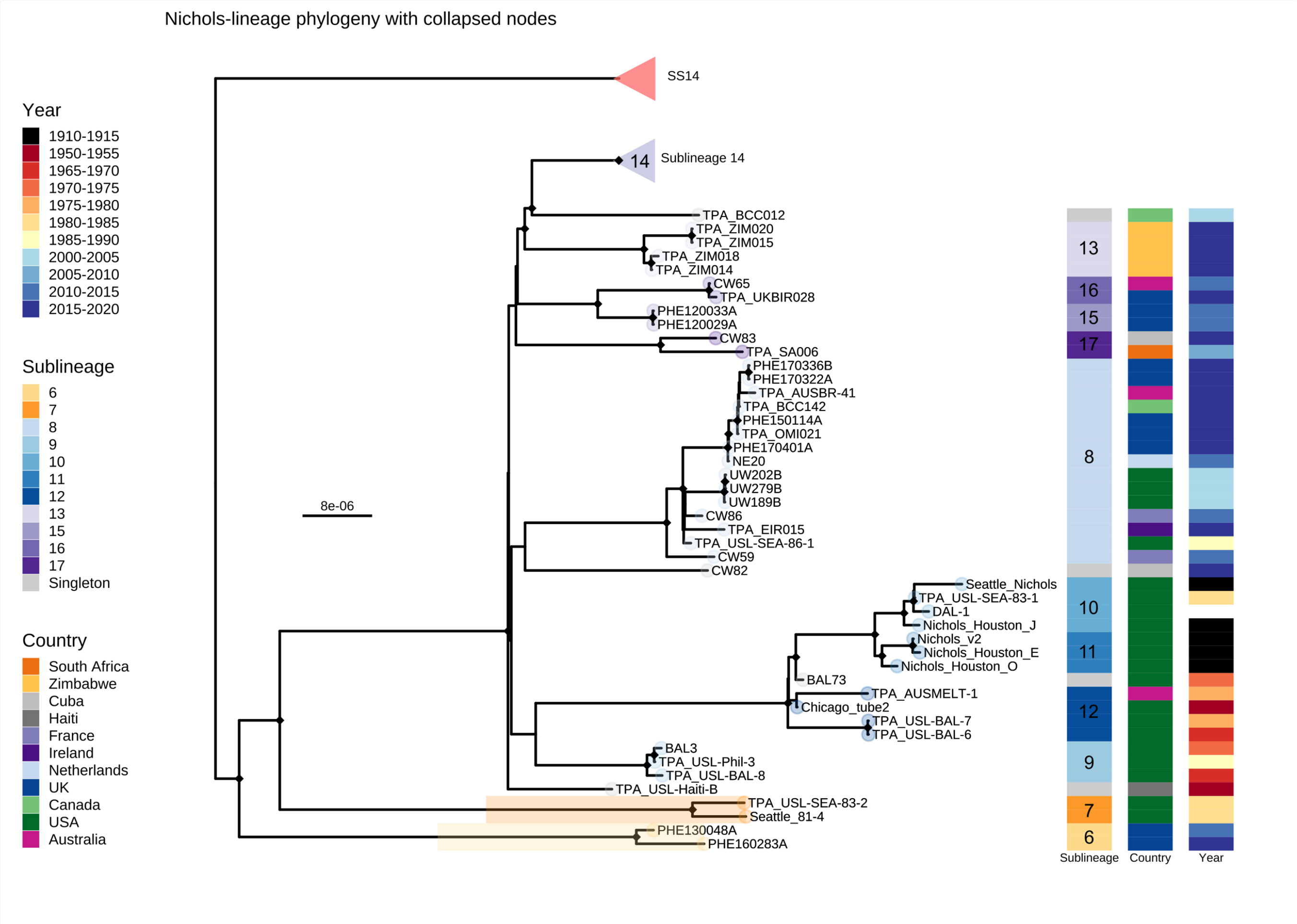


**Supplementary Figure 5. Subtree highlighting novel Nichols-lineage strains**. Recombination masked WGS phylogeny, showing the Nichols-lineage and sublineages. Tip points are coloured by sublineage, and coloured strips show sublineage and country. Shaded boxes highlight basal Nichols-lineage outgroup sublineages 6 and 7. The large clonal sublineage 14 has been collapsed to enable clearer visualization of the remaining taxa. The pink triangle indicates collapsed SS14-lineage, blue triangle indicates the collapsed sublineage 14.

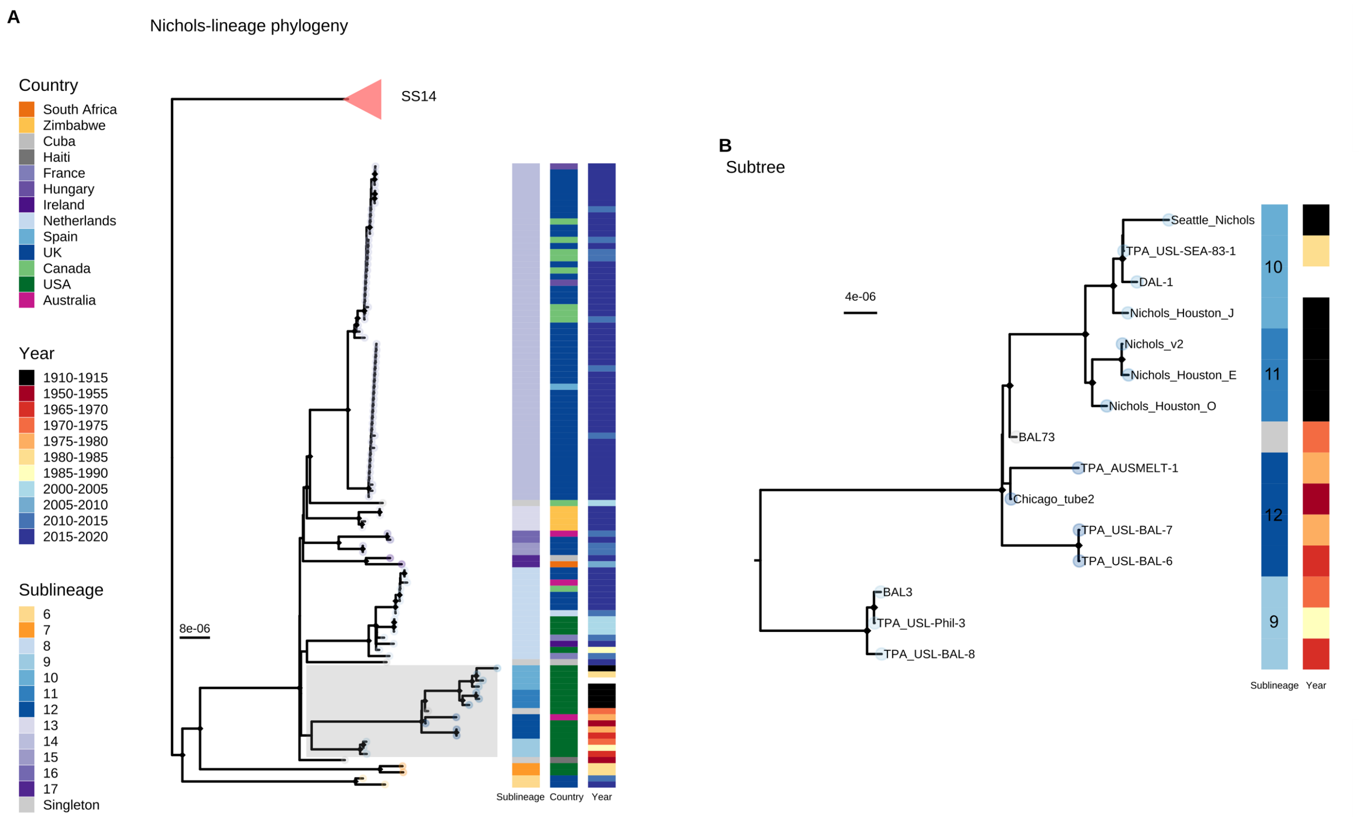


**Supplementary Figure 6. Commonly used Nichols Reference genomes form a monophyletic clade unrelated to contemporary clinical strains.** A- Recombination masked WGS phylogeny, showing the Nichols-lineage and sublineages. Shaded grey box shows a monophyletic clade containing commonly used reference genomes as well as genetically related strains. Tip points are coloured by sublineage, and coloured strips show sublineage and country. Pink triangle indicates collapsed SS14-lineage. B- Expanded view of a seemingly extinct clade containing common reference strains including Nichols_v2, DAL-1 and Seattle_Nichols. The most recent sample closely related to the reference strains (TPA_USL-SEA-83-1) was collected in 1983, while the latest sample for the entire clade (TPA_USL-Phil-3) was collected in 1987. The provenance of the sample originally used for sequencing the DAL-1 genome is uncertain, but in the literature the original isolation was in 1988. The placement of both DAL-1 and TPA_USL-SEA-83-1 within the diversity of Nichols-1912 derivatives suggests the possibility of the samples being mislabeled in the handling laboratories.

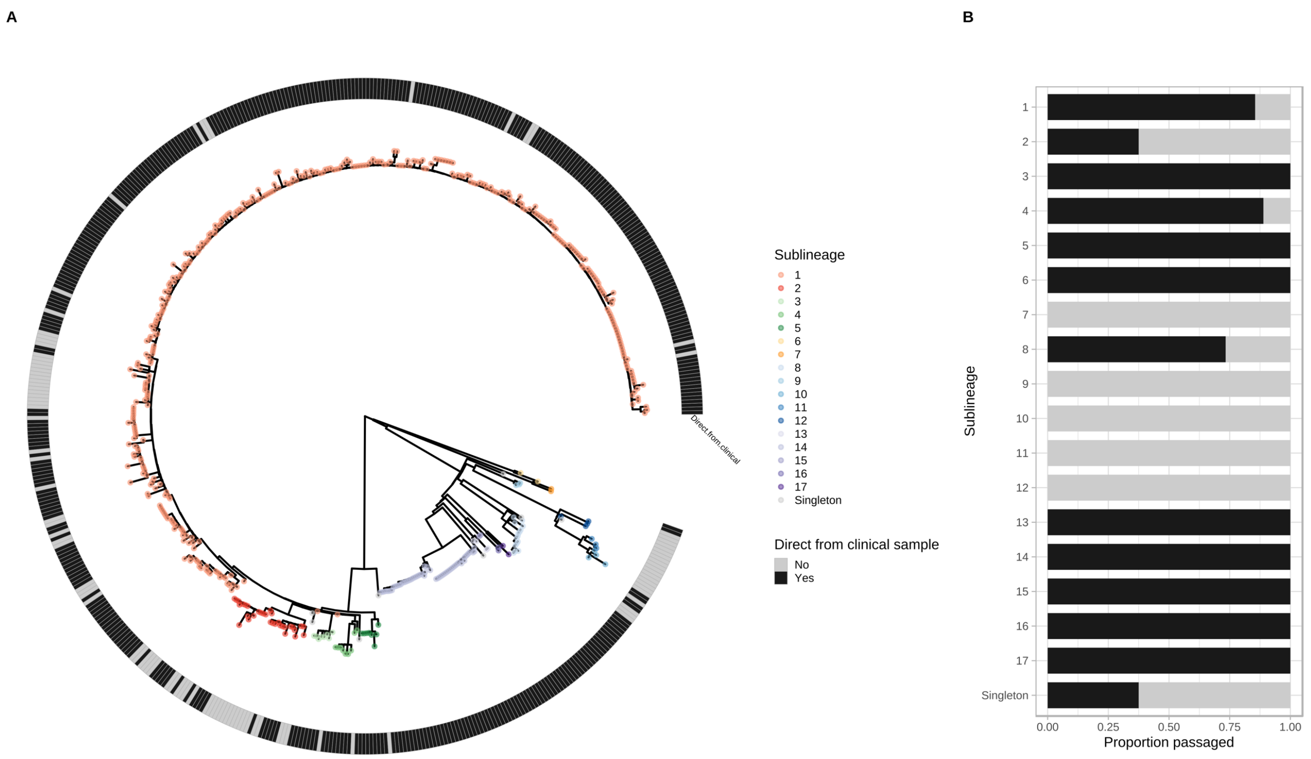


**Supplementary Figure 7. Finescale analysis of 528 high quality TPA genomes and sublineages, showing distribution of samples sequenced directly from clinical samples and those passaged in rabbit model.** A – Whole genome phylogeny showing distribution of samples sequenced directly from clinical sample or rabbit-passaged. B – Distribution of samples sequenced directly from clinical sample and rabbit-passaged samples according to sublineage. Samples passaged in rabbits are distributed throughout the global TPA phylogeny, and present in 9/17 sublineages. Older samples from before 2000 were isolated via rabbit passage, and dominate extinct clusters, as well as clustering close to the most recent common ancestor of contemporary sublineages such as SS14 sublineage 1.

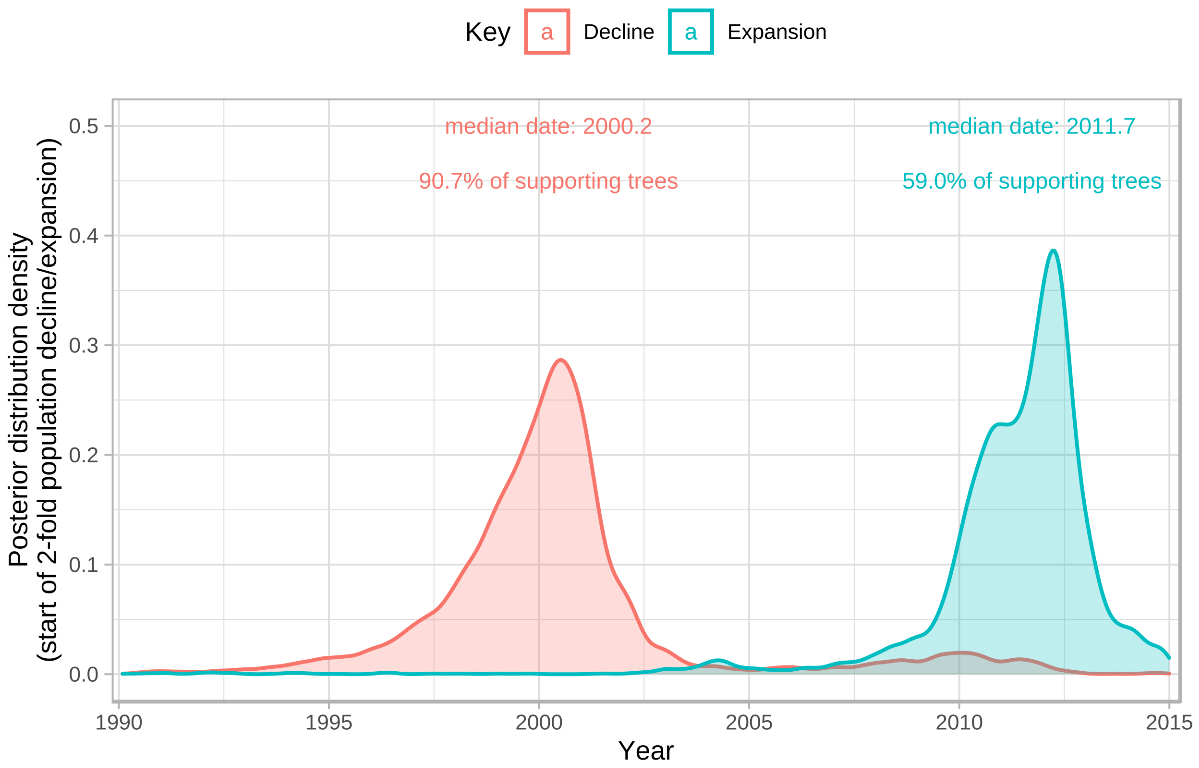


**Supplementary Figure 8. Bayesian Skyline analysis of population decline and expansion start dates.** Plots show posterior distribution of supporting trees for the start of either a 2-fold decline (pink) or expansion (blue) using a scanning approach within a window of 1990-2015. Analysis provides strong support for a population bottleneck in or around 2000, and moderate support for a subsequent expansion after 2010. Population changes are scaled to the population size averaged over the starting period for each tree. Therefore, if a particular tree already exhibited a decline near the starting timepoint, this may mean this tree does not show expansion, resulting in reduced overall support for expansion.

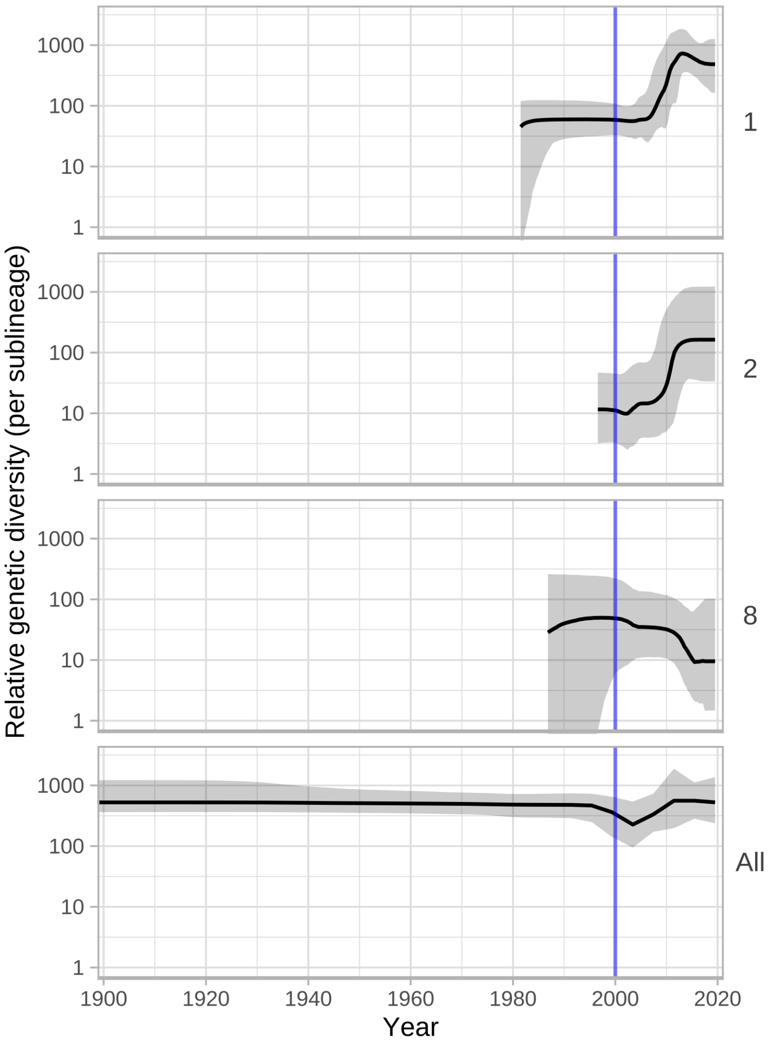


**Supplementary Figure 9. Bayesian Skyline analysis of sublineages.** Plots show population expansions occurring during the early 2000s for all sublineages with >15 samples apart from sublineage 14. Sublineage 14, which had low temporal signal, did not converge after multiple BEAST runs. Shows Skyline plots of sublineages 1, 2, 8 and plot for all samples from Figure 5.

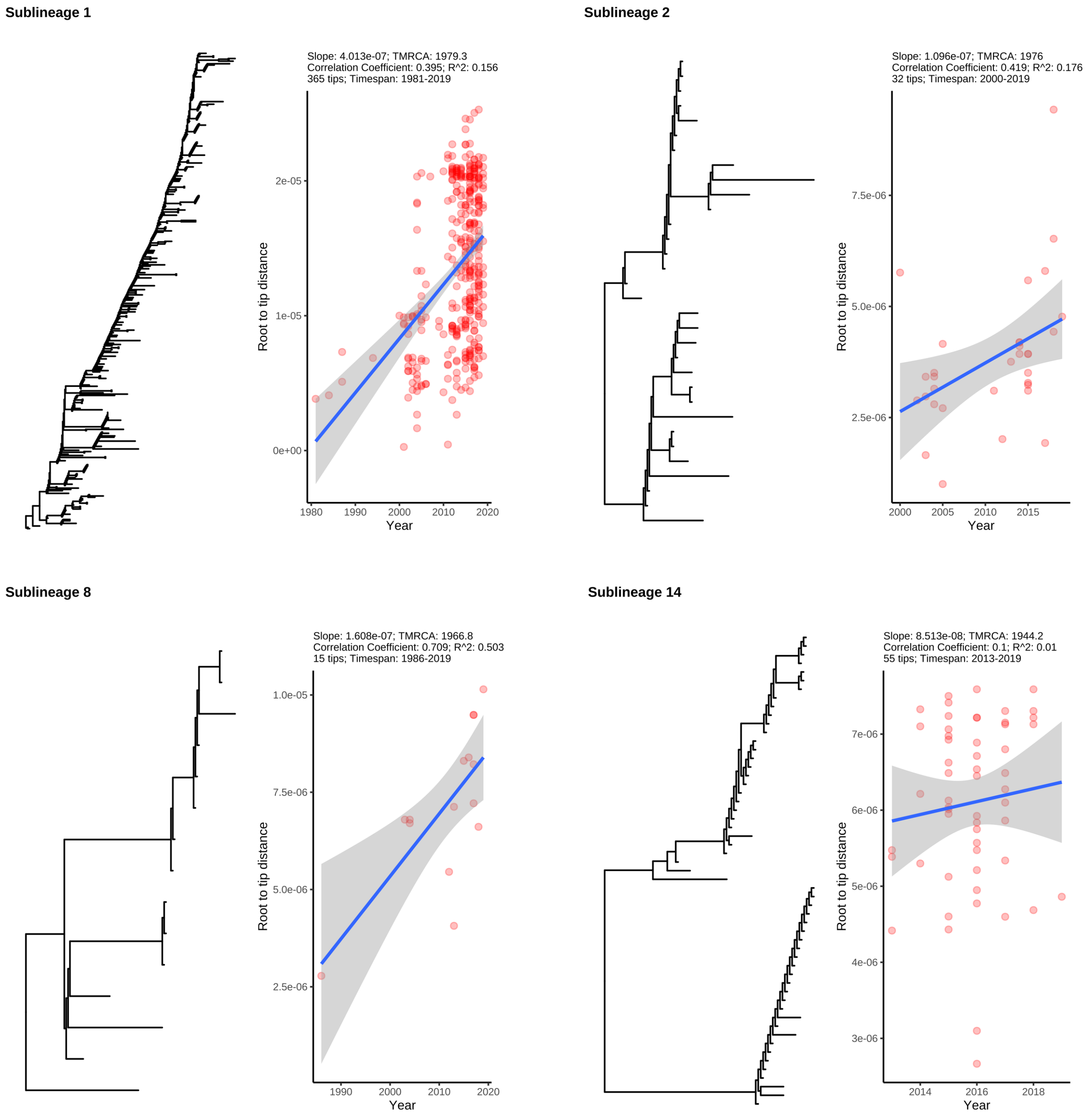


**Supplementary Figure 10. Subtrees of major sublineages, with corresponding root-to-tip distance plots.** All subtrees showed some evidence of temporal signal, but this was very weak for the recently emerged sublineage 14. Graphs are annotated with slope and time to most recent common ancestor (TMRCA) inferred directly from the maximum likelihood subtree, not BEAST.

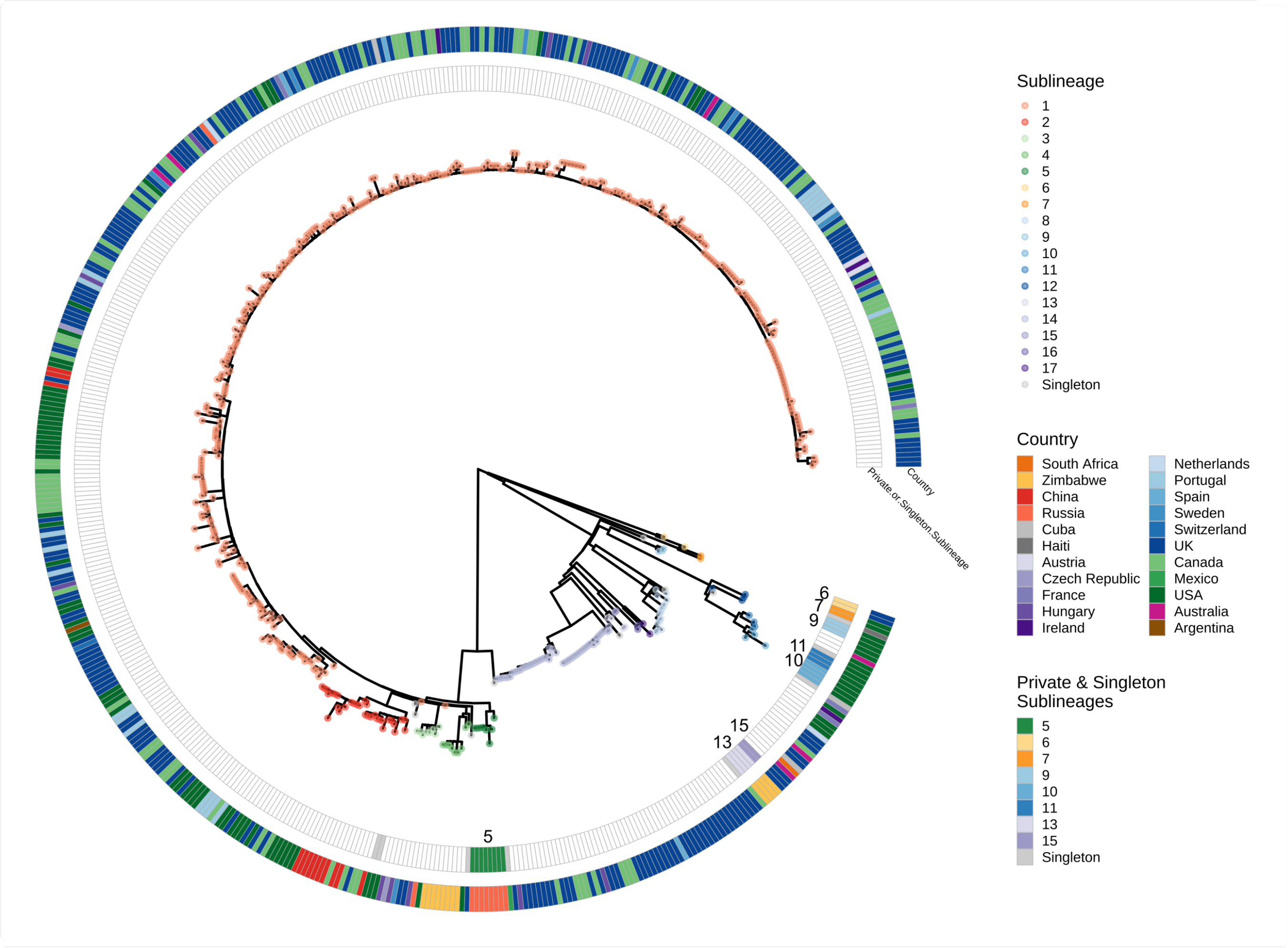


**Supplementary Figure 11. Finescale analysis of 528 high quality TPA genomes and sublineages, highlighting private and singleton sublineages.** Private and singleton sublineages are nested within the existing diversity of the TPA phylogeny. Tip points indicate sublineage, coloured tracks highlight singletons or private sublineages (with corresponding sublineage number), and country.

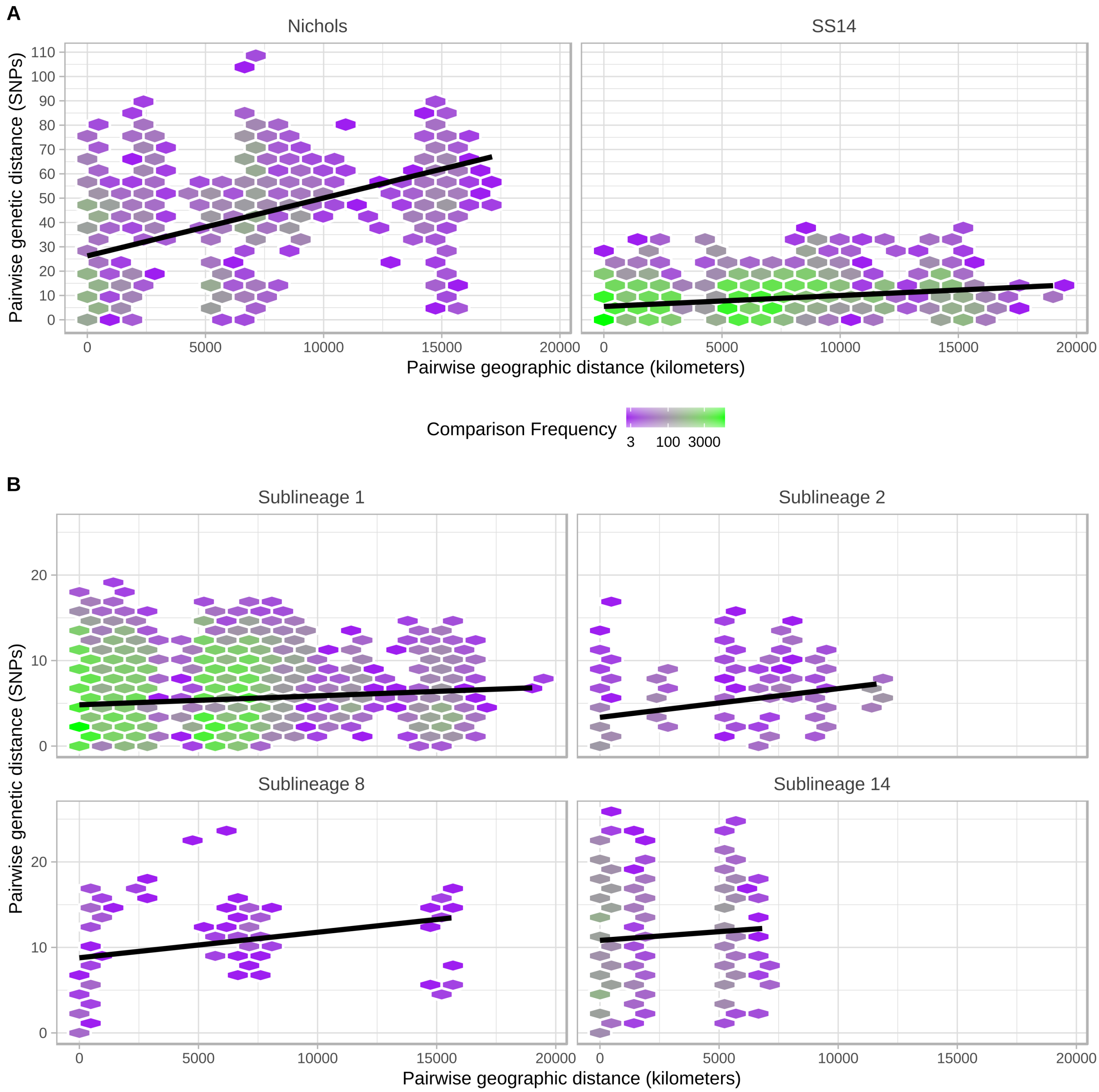


**Supplementary Figure 12. Effect of geographic distance on genetic distance.** A- Pairwise comparison of genetic distance (SNPs) and geographic distance (km; calculated using country centroids) within Nichols- and SS14-lineages, including linear regression (95% CI not visible). B- Pairwise comparison of genetic distance (SNPs) and geographic distance (km; calculated using country centroids) within the four major multi-country sublineages (SS14: 1, 2; Nichols: 8, 14). Includes linear regression (95% CI not visible).

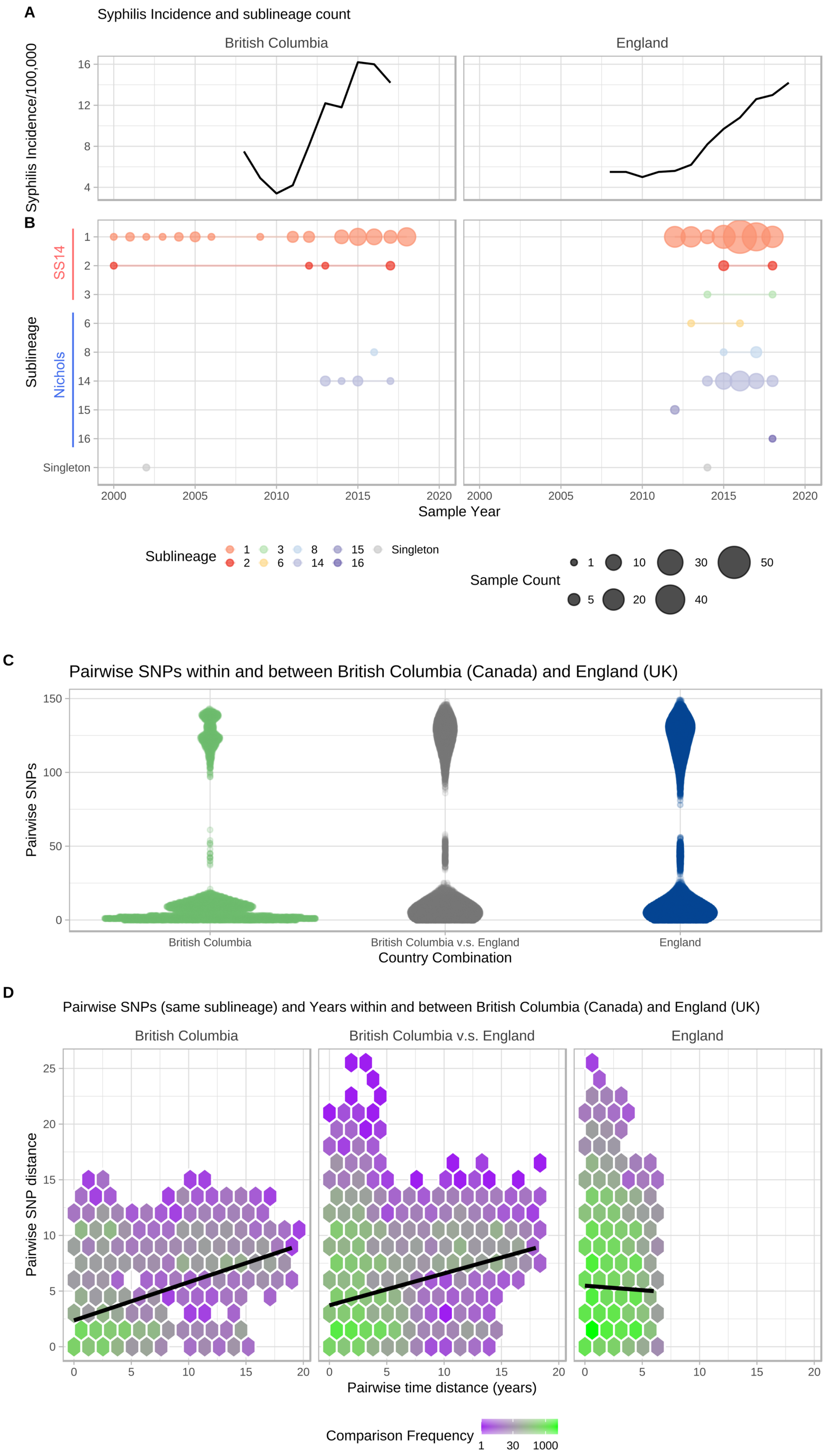
A
Syphilis Incidence and sublineage count
British Columbia
England
Syphilis Incidence/100,000
B
Sublineage
SS14
Nichols
Singleton
Sample Year
Sublineage
Singleton
Sample Count
C
Pairwise SNPs within and between British Columbia (Canada) and England (UK)
Pairwise SNPs
British Columbia
British Columbia v.s. England
England
Country Combination
D
Pairwise SNPs (same sublineage) and Years within and between British Columbia (Canada) and England (UK)
British Columbia
British Columbia v.s. England
England
Pairwise SNP distance
Pairwise time distance (years)
Comparison Frequency

**Supplementary Figure 13. Sharing of sublineages and closely related strains within and between British Columbia (Canada) and England (UK).** A- Syphilis incidence per 100,000 population by year for British Columbia, (Canada) and England (UK) using currently published data. B- TPA sublineage counts for each year, using high quality genomes from British Columbia (n=84) and England (n=240). British Columbia samples collected from 2000-2018, English samples collected from 2012-2018. C- Pairwise comparison of SNP distance distributions from samples within and between British Columbia and England. D- Comparison of SNP distance and temporal distance within and between British Columbia and England. The plot is divided into hexagonal bins, with the colour of each hexagon representing the number of comparisons (white=none, purple=few, green=many, see scale). Linear regression lines also shown (95% CI not visible).

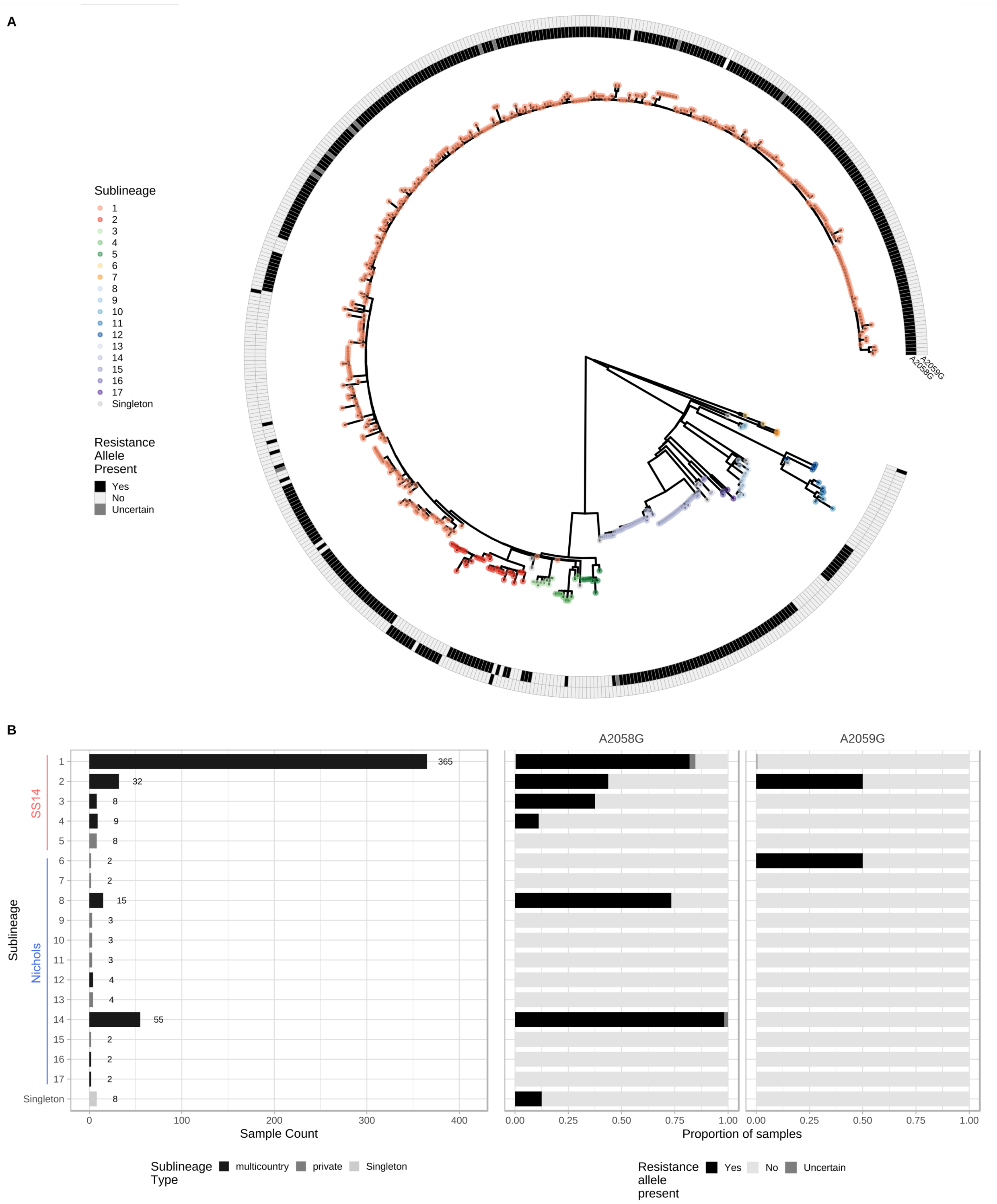


**Supplementary Figure 14. Multicountry sublineages are broadly macrolide resistant.** A- Whole genome phylogeny showing distribution of macrolide resistance conferring SNPs (A2058G and A2059G). B- Distribution of macrolide resistance SNPs by sublineage, indicating number of samples per sublineage, and sublineage type. Note that while the common A2058G mutation was found in six sublineages (both Nichols- and SS14-lineages), we also found the less common A2059G in both SS14-lineage (sublineages 1, 2) and Nichols-lineage (sublineage 6).

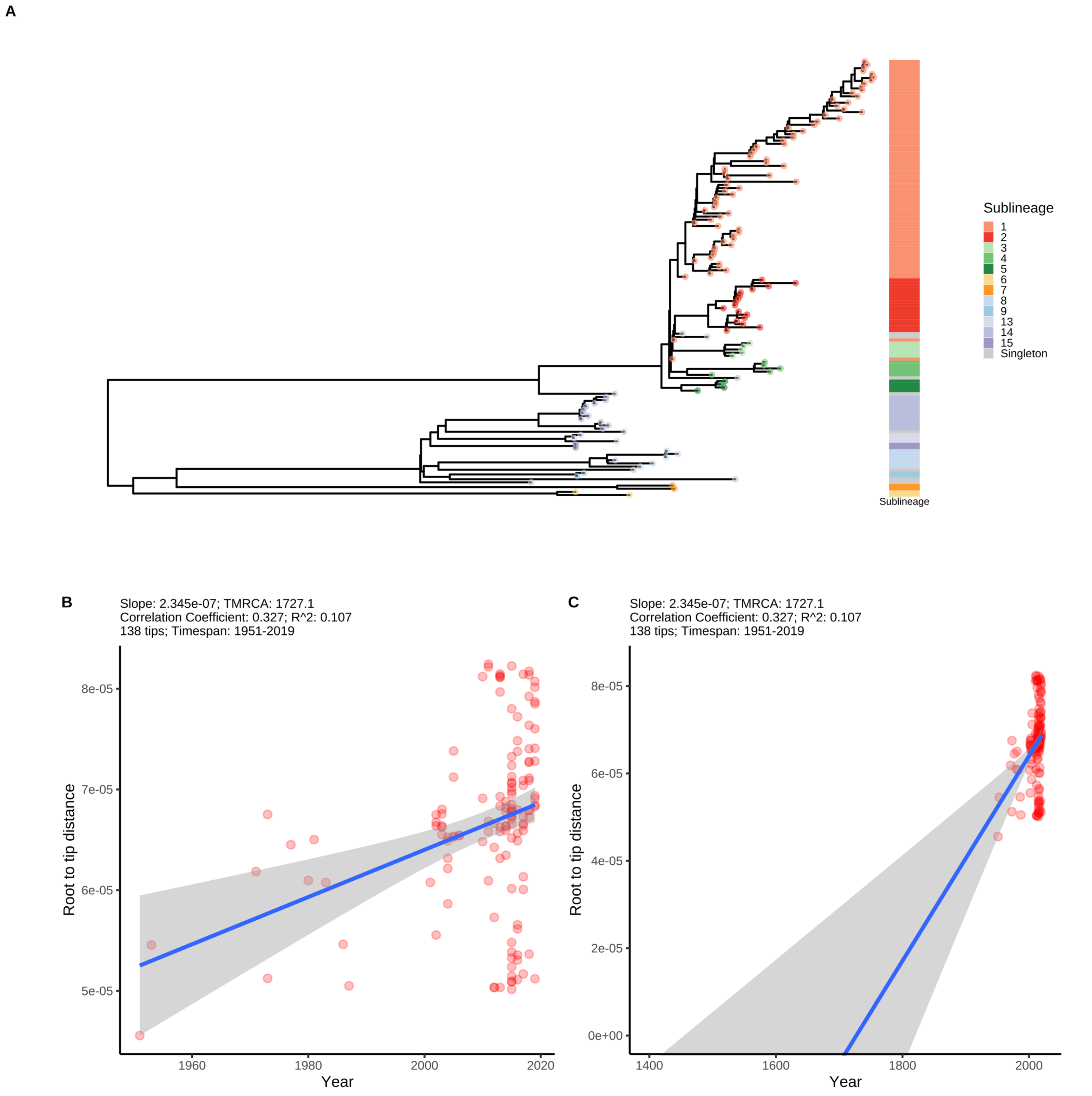


**Supplementary Figure 15. Maximum Likelihood phylogeny of 138 representatively subsampled genomes.** A- Maximum likelihood phylogeny of 138 genomes randomly sampled to be representative of sublineage and country. B- Scatterplot showing root-to-tip distance against collection date, illustrating temporal signal in the dataset. C- Expanded version of B, showing regressed x-intercept.

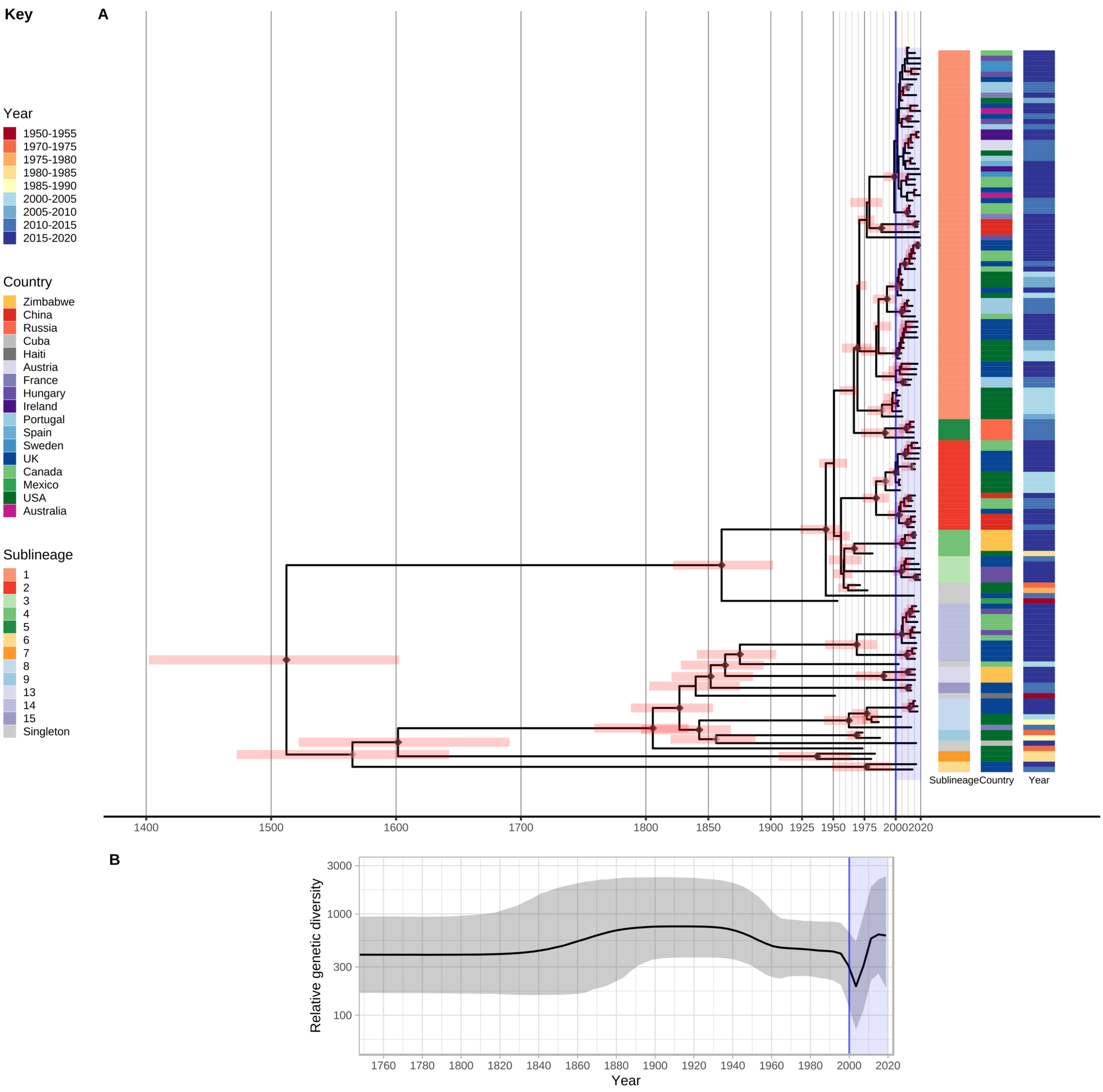


**Supplementary Figure 16. Bayesian maximum credibility phylogeny of 138 representative genomes shows population contraction during the 1990s, followed by rapid expansion from the early 2000s onwards.** A- Time-scaled phylogeny of 138 genomes randomly sampled to be representative of sublineage, country and collection year. Coloured tracks indicate sublineage, country and collection year. Node points are shaded according to posterior support (black ≥96%, dark grey >91%, light grey >80%). Red bars on nodes indicate 95% Highest Posterior Density intervals. Blue line and shaded area highlights post-2000 expansion of lineages. B- Bayesian Skyline plot shows decline of effective population size after the second world war, flattening in the 1960s, followed by a sharp decline and rapid reemergence during the 1990s and 2000s.

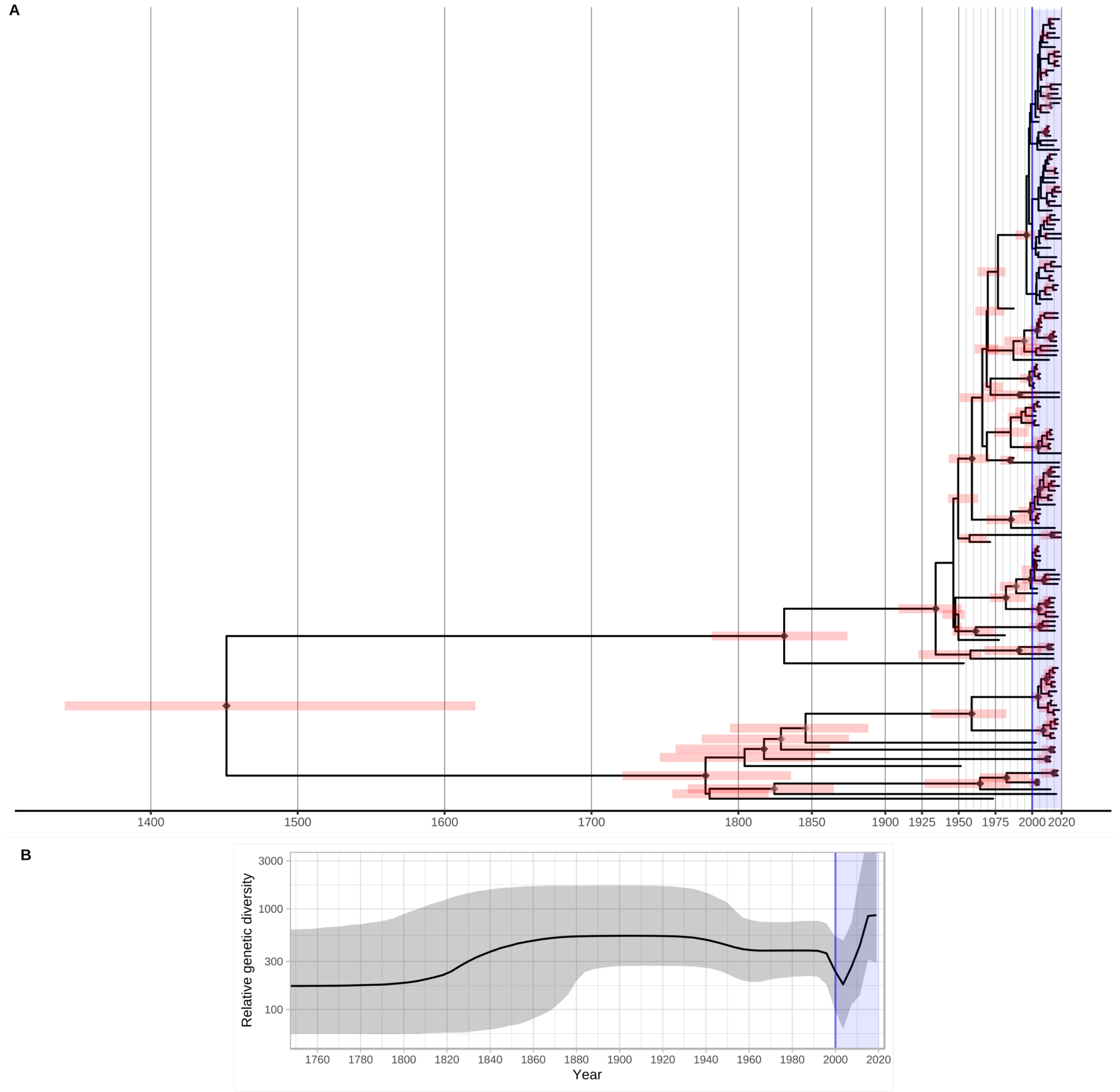


**Supplementary Figure 17. Secondary BEAST analysis of 168 separately subsampled representative genomes.** A- Time-scaled phylogeny of 168 genomes randomly sampled to be representative of sublineage, country and collection year. Node points are shaded according to posterior support (black ≥96%, dark grey >91%, light grey >80%). Red bars on nodes indicate 95% Highest Posterior Density intervals. Blue line and shaded area highlights post-2000 expansion of lineages. B- Bayesian Skyline plot shows decline of effective population size after the second world war, flattening in the 1960s, followed by a sharp decline and reemergence during the 1990s and 2000s, indicative of a sharp bottleneck.

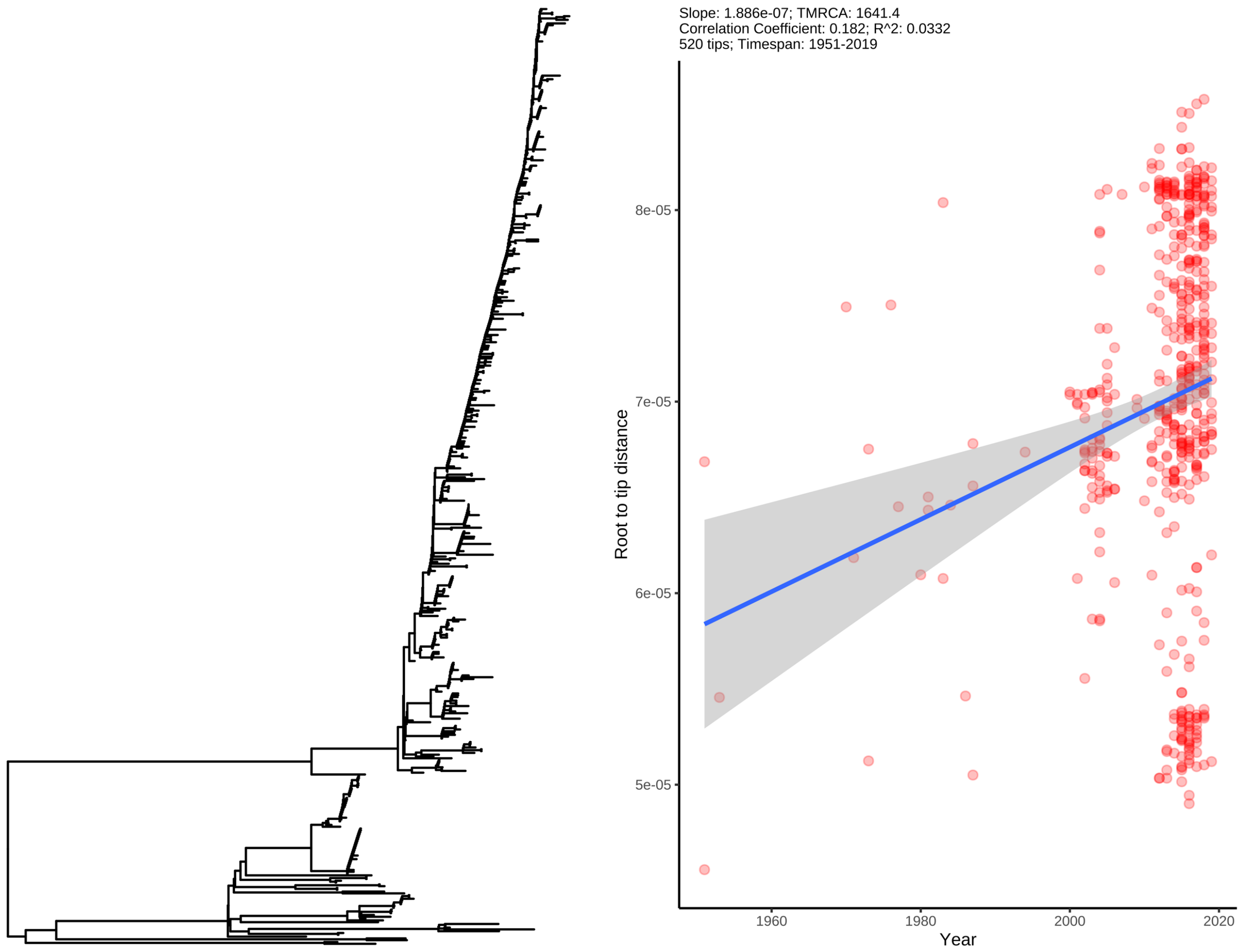


**Supplementary Figure 18. Maximum likelihood tree of 520 TPA genomes with minimal passage and robust collection dates, with corresponding root-to-tip distance plot.** Within the full tree, the temporal signal was weaker than in our subsampled dataset, but still plausible, given our prior analyses. Graphs are annotated with slope and time to most recent common ancestor (TMRCA) inferred directly from the maximum likelihood subtree, not BEAST.

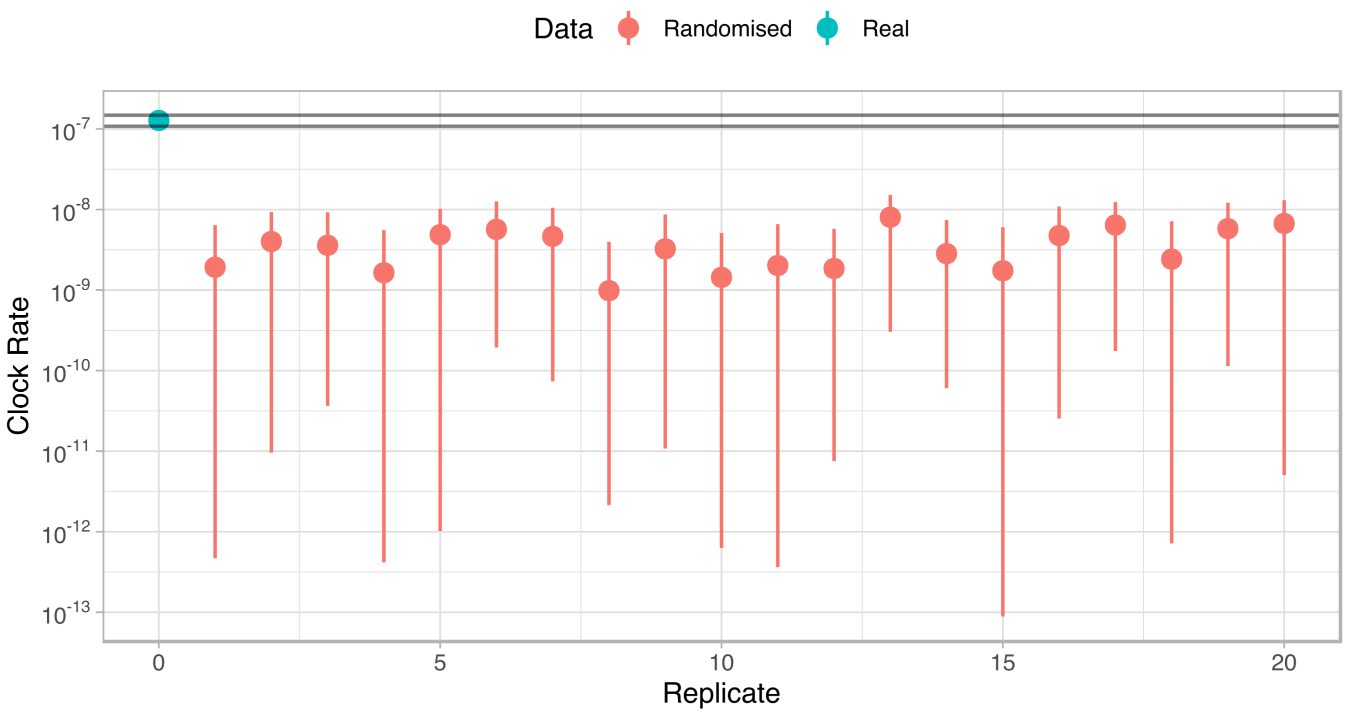


**Supplementary Figure 19. Date Randomisation Test for full BEAST2 dataset confirms the temporal signal in the true dataset compared to 20 resampled datasets with randomly reassigned tipdates.** The median clock rate for the real dataset was 1.27 x$10^{-7}$, while all randomly assigned datasets gave substantially lower clock rates; the highest median clock rate for the randomized datasets was 8.02 x$10^{-9}$. Real sample (blue), randomized samples (pink).

**Supplementary Data 1.** Metadata and read accessions for all samples included in this study

**Supplementary Data 2.** Genomic regions masked due through prefiltering or recombination analysis.
